# Peak alpha frequency is associated with pain severity in Long COVID patients with new-onset chronic pain

**DOI:** 10.64898/2026.02.16.26346388

**Authors:** Bárbara Silva-Passadouro, Omar Khoja, Alexander J. Casson, Ioannis Delis, Christopher Brown, Manoj Sivan

**Affiliations:** Leeds Institute of Rheumatology and Musculoskeletal Medicine, School of Medicine, University of Leeds, Leeds, UK; Department of Electrical and Electronic Engineering, University of Manchester, Manchester, UK; School of Biomedical Sciences, Faculty of Biological Sciences, University of Leeds, Leeds, UK; School of Electrical and Computer Engineering, National Technical University of Athens, Athens, Greece; Department of Psychology, Institute of Population Health, University of Liverpool, Liverpool, UK; National Demonstration Centre in Rehabilitation Medicine, Leeds Teaching Hospitals NHS Trust, Leeds, UK

**Keywords:** Electroencephalography, Post-Covid Syndrome (PCS), Post Acute Covid Syndrome (PACS), Peak alpha frequency, Pain, Resting-state

## Abstract

New-onset chronic pain is a common and debilitating symptom of Long COVID (LC) that remains not fully understood in terms of pathophysiology and therapeutic targets. A growing body of evidence in chronic pain syndromes similar to LC demonstrates an association between EEG alpha oscillatory activity and the experience of pain, with clinical studies showing maladaptive changes in oscillatory activity, particularly a slowing of alpha activity. This study aims to investigate the association between EEG alpha oscillatory activity and pain perception in new-onset LC-chronic pain. We recruited 31 individuals (20 females) with a clinical diagnosis of LC reporting new-onset chronic pain and 31 healthy pain-free age-and sex-matched controls. Participants completed questionnaires regarding symptoms and psychological functioning prior to recording eyes-open resting-state EEG. Peak alpha frequency (PAF) and spectral power within the alpha band (8–13 Hz) were extracted from EEG signals. Lower PAF over the posterior scalp region was significantly associated with higher LC-chronic pain severity when controlling for age and depression. This observation was consistent across PAF estimation methods. PAF was significantly increased, particularly in the posterior region, in the moderate pain LC subgroup compared to both severe pain subgroup and controls, while alpha power did not differ between the three groups and was not associated with pain severity. Our findings highlight associations between PAF and pain symptoms in a new post-infection chronic pain syndrome. PAF can thus be explored as a potential biomarker and therapeutic target for EEG-based neuromodulation interventions in LC-chronic pain. These results may have implications for other similar chronic pain syndromes.

**Summary:** Lower resting-state EEG peak alpha frequency in posterior scalp region is associated with higher severity of new-onset Long COVID chronic pain.

## Introduction

Pain is a common and often debilitating symptom of Long COVID (LC), a multisystem post-infection syndrome experienced by over 400 million people globally [10]. New-onset musculoskeletal LC-chronic pain refers to pain of musculoskeletal origin that persists or arises following resolution of the acute phase of coronavirus disease-19 (COVID-19) infection and lasts longer than three months [62]. Chronic pain is consistently cited as one of the most commonly reported symptoms of LC, estimated to be present in at least 10 to 20% of LC cases [18,33,35,47,50], with prevalence rates going up to 50% in patients hospitalised due to COVID-19 illness [16,24,61].

The widespread distribution of pain typically observed in LC and its comorbid association with other central nervous system-related symptoms such as sleep disturbances, depression, fatigue and cognitive impairments, are suggestive of a nociplastic-type pain [6,17,19,36,45,49]. In fact, a significant proportion of LC patients fulfil the diagnostic criteria for Fibromyalgia Syndrome (FMS) [27,34,55,63], where nociplastic pain is typically present [38]. The lack of reliable diagnostic tools and effective biomarkers makes nociplastic pain difficult to identify and treat, which has important clinical implications for LC, where the coexistence of mixed pain types adds further complexity to clinical management.

Emerging evidence suggests that neurophysiological markers, such as oscillatory cortical neural activity measured via electroencephalography (EEG), may offer valuable insights into the neural basis of chronic pain. There has been particular interest directed toward changes in alpha oscillatory activity (8–13 Hz) as a potential biomarker of chronic pain [52,68]. For instance, studies have reported a slowing of EEG alpha activity, as indexed by decreased peak alpha frequency (PAF), in individuals with chronic pain, including those with FMS, compared to pain-free controls [40,54,65]. PAF slowing has been consistently associated with heightened pain sensitivity and sustained pain in experimental models [20,21], with further evidence to support its predictive value for post-operative pain severity [44]. There is also evidence of reduced alpha power in FMS patients [28,41,64], although this finding does not appear to be fully consistent with observations in conditions associated with neuropathic pain [5,54,67]. Given the overlap in clinical features, it may be hypothesised that LC-chronic pain, which phenotypically resembles FMS, is associated with alterations in EEG alpha activity comparable to those observed in FMS [59].

This study aims to investigate EEG alpha oscillatory features of new-onset LC chronic pain. The data was derived from the Musculoskeletal Pain in Long COVID (MUSLOC) research project (clinicaltrials.gov NCT05358119), a prospective longitudinal study aiming to investigate the characteristics and mechanisms underlying LC-chronic pain in a sample of

LC patients with new-onset musculoskeletal chronic pain. Here, we present the findings of cross-sectional analysis of resting-state EEG data from the same cohort. A healthy pain-free control group was recruited separately for comparative analysis with patient data. We hypothesised that (1) lower PAF is associated with higher severity of new-onset LC chronic pain; (2) LC patients with new-onset chronic pain have lower PAF compared to pain-free controls; and (3) alpha band power is decreased in patients compared to controls.

## Methods

### Participants

Thirty-one adult patients with LC and new-onset chronic pain were recruited primarily from the Leeds Long COVID Community Rehabilitation Service, as well as through social media and patient support groups. Following initial interest, individuals were contacted to confirm eligibility and to explain the study aims and procedures. The criteria for inclusion in the study was: (1) aged 18 years or older; (2) tested positive for COVID-19 or reported COVID-19 symptoms that were confirmed by an independent clinician; (3) had a clinical diagnosis of LC as per the NICE guidelines definition [46]; (4) presented with new-onset musculoskeletal chronic pain as defined by the ICD-11 criteria [62] since COVID-19 infection. Potential participants were excluded if they presented with chronic pain prior to COVID-19 infection; were unable to read and understand English; had history of seizures or clinical diagnosis of epilepsy; or were unable to comply with study procedures.

The control group consisted of 31 age- and sex-matched healthy volunteers with no clinical history of seizures or epilepsy and not experiencing any pain. Healthy controls were recruited in a separate study (outside the MUSLOC study for patients) via social media advertisement and university mailings lists. The EEG equipment, recording environment, recording parameters, protocol, signal preprocessing and EEG feature extraction procedure were identical for patient group and healthy controls.

All participants provided written informed consent prior to any study procedure and were informed of their right to withdraw from the study at any time. Participation in the study was entirely voluntary. The MUSLOC project was registered on clinicaltrials.gov (NCT05358119) and approved by the London - Central Research Ethics Committee (REC), the Health Research Authority (HRA) and Health and Care Research Wales (HCRW) (Ref 21/PR/1377). Healthy participants were recruited under ethical approval from the University of Leeds’ Faculty of Biological Sciences ethics committee (BIOSCI 23-008). All procedures were conducted in accordance with the Declaration of Helsinki.

### Questionnaires

Patients completed the self-assessment questionnaires listed below prior to EEG data collection:

a. Demographic information form: to collect demographic data, including items in regard to participant’s age, sex, ethnicity, marital status, weight, height, pre-existing medical conditions, smoking status, COVID-19 related information (i.e. COVID-19 infection and vaccination history, hospitalisation due to COVID-19), employment category and status following COVID-19 pandemic and LC.
b. COVID-19 Yorkshire Rehabilitation Scale (C19-YRS): the first validated scale developed specifically for assessment and monitoring of patients with LC and used in LC clinics across the UK [60]. It consists of 15 items relative to severity of core symptoms of LC (including breathlessness, cough, swallowing, fatigue, continence, pain, cognition, anxiety, depression, post-traumatic stress disorder) and impact on daily functions (including communication, mobility, personal care, other activities of daily living, social role). Each item is scored on two numeric rating scales ranging from 0 to 10 relative to their symptoms pre- and post-COVID respectively. Items 1–10 capture symptom severity (scores 0–100) and items 11–15 capture functional disability (scores 0–50). The C19-YRS also measures the severity of other common LC symptoms (including palpitations, dizziness, weakness, sleep problems, fever, skin rash) and overall health (scores 0–10; score of 0 for worst health and 10 for best health imagined). The post-COVID pain item of the C19-YRS was used as pain severity score in the analysis for this study.
c. Patient Health Questionnaire 9 (PHQ-9): a screening, diagnostic and monitoring tool for depression based on the diagnostic criteria set by the Diagnostic and Statistical Manual of Mental Disorders [39]. It consists of nine items rated on a 4-category response scale (0 – Not at all; 1 – Several days; 2 – More than half the days; 3 – Nearly every day) that ask individuals to rate how often they have been bothered by symptoms of depression in the past two weeks. The level of severity of depression is assessed using the total scores (range: 0–27), with 10 being the standard cut-off score for major depression [23].

Healthy volunteers completed a demographic information form and the PHQ-9.

### EEG recordings and data acquisition

Resting-state EEG signals were collected using a BrainVision actiCHamp Plus amplifier system (Brain Products GmbH, Gilching, Germany) with active Ag/AgCl electrodes attached to a 64-channel ActiCAP (Brain Products GmbH, Wörthsee, Germany) positioned according to the 10–20 international electrode placement system. Participants were sat comfortably on a chair in a dim light room. They were instructed to remain relaxed and keep their eyes open whilst fixating on a cross on the wall approximately one meter from the chair. Two blocks of six minutes each of resting-state EEG signal were recorded for each participant. EEG signals were recorded at a sampling rate of 1000 Hz and electrode impedances were kept at 10 kΩ or below. All EEG electrodes were referenced to the FCz electrode. The ground electrode was placed at Fpz channel position.

### EEG preprocessing

Pre-processing and cleaning of raw EEG signals was implemented in MATLAB (ver. R2023b) custom scripts using the EEGLAB toolbox [11]. Data were filtered using zero-phase, linear phase finite impulse response (FIR) filters. A high-pass filter and low-pass filter were applied sequentially at frequencies 0.5 Hz and 35 Hz respectively, followed by attenuation of 50 Hz power line interference by notch filtering. After downsampling to 250 Hz, EEG data recorded from each participant were merged, resulting in one single 12-minute dataset per participant. Flat or excessively noisy channels were then removed, with a mean of 0.4 channels per participant (range: 0–4). The Artifact Subspace Reconstruction (ASR) algorithm was applied to automatically correct bad data periods with variance larger than the rejection threshold of 30 standard deviations (*k* parameter) relative to a clean portion of EEG signals (*clean_rawdata* plugin for EEGLAB) [9]. Other ASR parameters were set to default.

The data were then re-referenced to the common average. Independent component analysis (ICA) was computed using EEGLAB function *runica* [32], with the number of components limited to the effective data rank [11]; data were internally whitened using a principal component analysis (PCA)-based step prior to ICA optimisation [30,32][32]. Components generated by eye blinks, saccades, muscle activity and heart artifacts were identified with the support of ICLabel classification [51], with any components labelled as >90% non-brain artifact automatically flagged for rejection. Further rejection was performed manually through visual inspection based on component topographies, frequency distribution and time courses (median of total number of ICA components rejected = 9, range: 2–19, IQR: 6.25–11). Rejected channels were interpolated after ICA cleaning.

### EEG feature extraction

Power spectral density for each channel was estimated from the cleaned EEG signals using the Welch’s method (2-s segments, no overlap, Hanning window). The absolute spectral power of the alpha frequency band (8–13 Hz) was extracted using the trapezoidal integration method of estimation of the area under the spectral curve (MATLAB function *trapz*). The power of other canonical frequency bands was also extracted for exploratory supplementary analysis, with ranges set as follows: delta (2–4 Hz), theta (4–8 Hz), and beta (13–30 Hz).

Peak alpha frequency was derived using two different methods. Max-PAF was estimated as the frequency in the alpha range (8–13 Hz) that exhibits maximum Welch’s power spectral density. The maximum Welch’s power spectral density was also extracted and set as max-PAF power spectral density. The second method used the spectral parameterisation algorithm (parameters set to default) to decompose Welch’s power spectral data into periodic and aperiodic components (*specparam* toolbox, formerly *fooof*, version 1.1.0; Python wrapper version 1.1.0) [14]. This approach accounts for the contribution of aperiodic EEG activity, characterised by a 1/f-like distribution, which co-occurs with periodic activity and can confound the estimation of true oscillatory peaks within canonical frequency bands. For instance, changes or interindividual variation in PAF or power at a specific frequency band can be conflated with differences in aperiodic activity, thus leading to misinterpretation of underlying physiological phenomena [3]. The *specparam* algorithm iteratively fits Gaussian curves to model each oscillatory peak in the aperiodic-adjusted power spectrum, derived by subtracting the estimated 1/f aperiodic component. The algorithm outputs parameters that define the aperiodic component (exponent and offset), the putative periodic component (centre frequency, power and bandwidth for each fitted Gaussian) and overall model fit (R^2^ and error). For the present study, aperiodic-adjusted PAF was set as the frequency with the strongest Gaussian peak identified within the alpha range. Power spectral density of the aperiodic-adjusted PAF was also extracted from model outputs. Channels for which spectral parameterisation did not detect an alpha oscillatory peak were identified for each participant and excluded from max-PAF estimation for that participant (mean of 3.4 channels per participant, range: 0–24). We excluded channels with *specparam* model fit below <0.9 R^2^ from analysis as suggested by Gyurkovics *et al*. [26].

Peak alpha frequency and power estimates were median averaged across respective channels to extract global (all electrodes) and regional PAF for each participant. Three regions of interest were defined for regional analysis: anterior region (comprising electrodes Fp1, Fp2, AF7, AF3, AFz, AF4, AF8, F7, F5, F3, F1, Fz, F2, F4, F6 and F8); central region (comprising electrodes FT9, FT7, FC5, FC3, FC1, FC2, FC4, FC6, FT8, FT10, T7, C5, C3, C1, Cz, C2, C4, C6, T8, TP9, TP7, CP5, CP3, CP1, CPz, CP2, CP4, CP6, TP8 and TP10); and posterior region (comprising electrodes P7, P5, P3, P1, Pz, P2, P4, P6, P8, PO7, PO3, POz, PO4, PO8, O1, Oz and O2).

### Statistical analysis

Statistical analysis was conducted in RStudio (version 2024.12.1). Data are presented as mean ± standard deviation unless otherwise indicated. Statistical significance for all tests was set at α = 0.05. Participants with missing or incomplete questionnaires were excluded from the analysis relative to that specific questionnaire.

Spearman’s rank correlation was calculated between max-PAF and aperiodic-adjusted PAF estimates derived from each region to assess the robustness and consistency of the estimates across PAF extraction methods.

### Primary aim 1. Association between PAF and pain severity in LC patients with new-onset chronic pain

The association between pain symptoms and EEG PAF was investigated using multiple linear regression analysis. Peak alpha frequency derived from the two estimation methods (max-PAF and aperiodic-adjusted PAF) was entered as a predictor in separate models, with C19-YRS pain severity subscale scores as dependent variable. Global PAF estimates and estimates derived from each region (anterior, central and posterior) were tested separately. Depression and older age have been linked with decreased PAF [25,56,58,69]. Hence, we adjusted for the potential confounding effects of age and depression (as self-reported in the PHQ-9) in our analysis. All model assumptions were examined through model diagnostic plots including residual plots and Q-Q plots. The models were specified as:

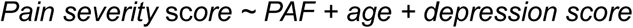

P values from the core adjusted models (derived from the formula above) were corrected for multiple testing using false discovery rate (FDR) correction. FDR correction was applied across models derived from global and all regional estimates, separately for each family of models relevant to the different PAF estimation methods. For transparency, we also present the outputs of simple regression models (i.e. *Pain severity score ∼ PAF*) and models including each of the confounding variables individually to show the effects of introducing each to the adjusted model in Supplementary Materials.

Significant PAF-pain associations were followed up with further analysis to assess the symptom-specificity of the findings. For that purpose, the severity of other neurological symptoms of LC (i.e. fatigue and cognitive impairment subscales of the C19-YRS), overall LC symptom severity, duration of pain symptoms and number of painful sites (pain widespreadness) were included as dependent variables in separate models. Additional exploratory analysis was performed with power of the strongest peak within the alpha range (PAF power spectral density) as independent variable, to test whether PAF power spectral density independently predicts pain.

### Primary aim 2. Comparative analysis of PAF between LC patient and healthy pain-free control groups

All variables were tested for normal distribution (using Shapiro-Wilk test, Q-Q plots and histograms) and equality of variances (using Levene’s test) prior to between-groups analysis. If found non-normally distributed, the channel-averaged EEG feature values were transformed using logarithmic transformation (log10).

Between-group differences in PAF were tested with multiple linear regression models including a 2-level group factor (patients versus controls). A secondary subgroup analysis was conducted by further stratifying patients based on pain severity. Patients were classified into moderate and severe pain subgroups based on their C19-YRS pain subscale scores.

The median was used as cut-off value [22,66], with individuals in the moderate pain group reporting pain severity scores below the median and individuals in the severe pain group reporting pain scores equal or above the median. The group factor for the subgroup analysis models consisted of 3-levels: moderate pain group, severe pain group and control group.

Peak alpha frequency derived from the two estimation methods (max-PAF and aperiodic-adjusted PAF) was entered as dependent variable in separate models. Additionally, global PAF estimates and estimates derived from each region (anterior, central and posterior) were tested separately. The models were specified as:

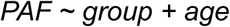

Each model was followed by an omnibus F-test of the coefficients, which allowed to determine the overall effect of group. Significant effects of group were followed by Tukey’s adjusted post-hoc pairwise comparisons.

Furthermore, we conducted exploratory non-parametric cluster-based permutation tests on electrode-wise data to confirm the findings of significant between-group differences from regional analysis. This approach allows for a more precise localisation of group differences across the scalp without predefining regions of interest and while controlling for multiple comparisons. Cluster-permutation testing was conducted using Fieldtrip (version 20230118) [48]. First, the dependent variable was regressed on age, with residuals then used as the input for cluster analysis. Channel neighbours were identified using the function *ft_prepare_neighbours* based on spatial proximity between electrodes (minimum neighbourhood distance parameter set to default), with a minimum of two neighbouring electrodes for cluster formation. The analysis was implemented with 10,000 permutations to detect significant spatial clusters across electrodes (two-tailed, cluster-forming threshold and cluster significance threshold set to α = 0.05).

### Primary aim 3. Comparative analysis of alpha band power between LC patient and healthy pain-free control groups

Comparative analysis of alpha band power between groups followed the methods described for primary aim 2, but with alpha power estimates as dependent variable.

#### Additional exploratory analysis (Supplementary Materials)

In addition to the prespecified primary analyses, we conducted a series of exploratory and confirmatory analyses to further test EEG alpha oscillatory activity patterns associated in LC-chronic pain. This included: (a) testing association between EEG alpha features and pain within LC patient group accounting for effects of centrally acting drugs; (b) sensitivity analysis of comparison between LC patients and controls accounting for the potential confounding effects of depression; (c) testing association between absolute spectral power of other canonical EEG frequency bands (delta, theta and beta) and pain severity within patient group; and (d) comparative analysis of absolute spectral power of other canonical EEG frequency bands between LC patients and controls.

No adjustments were made for multiple comparisons given the exploratory nature of these analyses. All methods and results related to these analyses are reported in the Supplementary Materials.

## Results

### Demographics and symptom severity

Table 1 presents a summary of participant demographics for both the LC patient group and the healthy pain-free control group, and Table 2 shows a summary of clinical characteristics and severity of neurological and psychological symptoms of LC based on the C19-YRS for the patient group. A full list of all the symptoms reported on the C19-YRS, including pre- and post-COVID scores, can be found in Supplementary Materials Tables S1.

**Table 1.** Summary of demographic and psychological variables for the patient and healthy pain-free control groups.

|  | Long COVID patient group<br>(n = 31) | Healthy pain-free control group<br>(n = 31) |
| --- | --- | --- |
| **Demographic characteristics** |  |  |
| Age, mean (SD), years | 46.4 (11.1) | 46.6 (11.0) |
| Sex, n (%) |  |  |
| female | 20 (64.5) | 20 (64.5) |
| male | 11(35.5) | 11(35.5) |
| Ethnicity, n (%) |  |  |
| white | 26 (83.9) | 23 (74.2) |
| asian | 5 (16.1) | 7 (22.6) |
| black | 0 (0.0) | 0.0 (0.0) |
| mixed | 0 (0.0) | 0.0 (0.0) |
| other | 0 (0.0) | 1 (3.2) |
| BMI, mean (SD), kg/m<sup>2</sup> | 31.4 (9.5) (missing: 6) | 24.4 (4.0) (missing: 1) |
| Smoking status, n (%) |  |  |
| none/former | 27 (87.1) | 31 (100.0) |
| active | 1 (3.2) (missing: 3) | 0 (0.0) |
| **Psychological variables** |  |  |
| PHQ-9 scores, mean (SD) | 12.8 (5.3) (missing: 6) | 2.8 (2.8) |

**Table 2.**
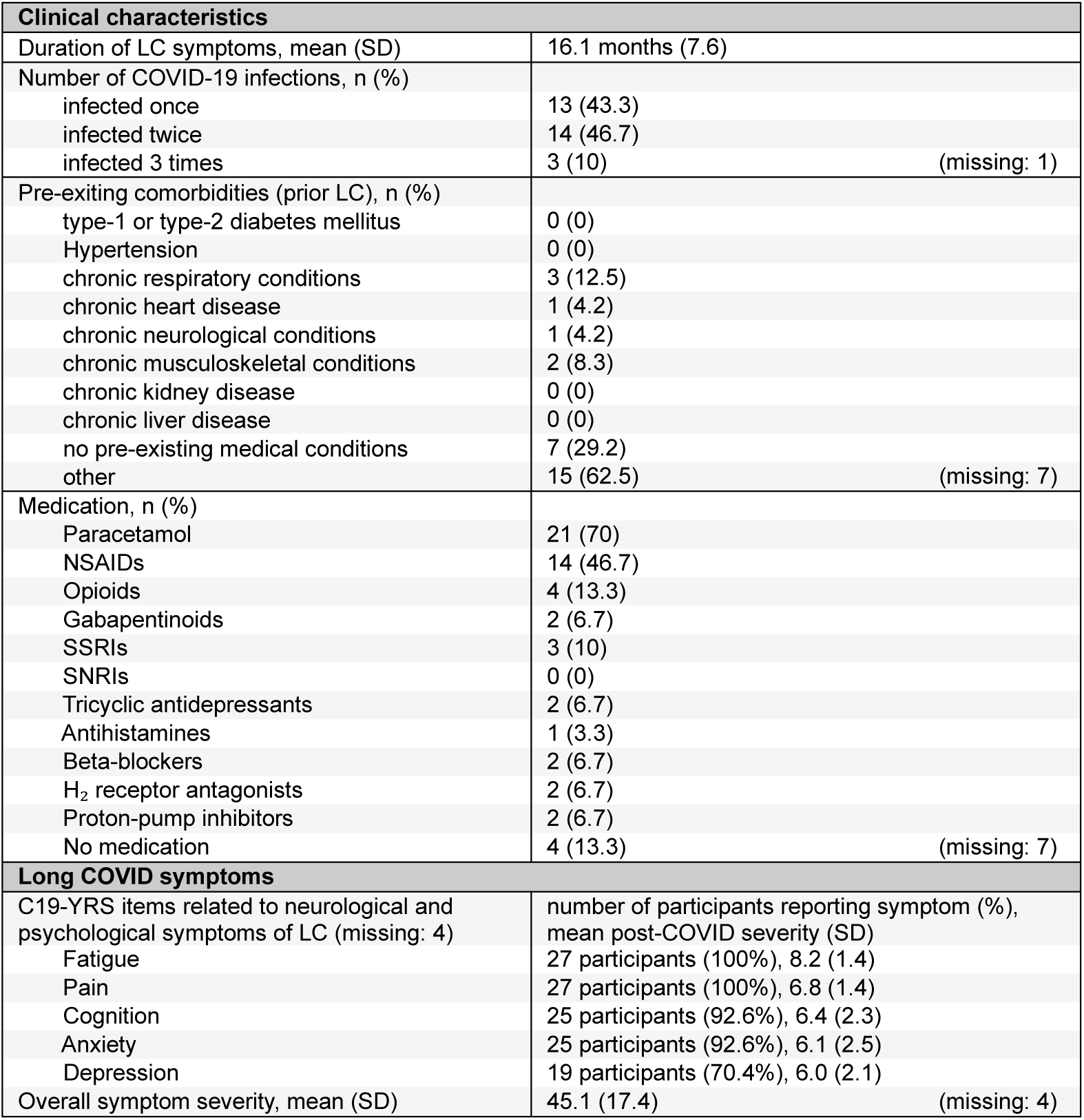
Summary of clinical characteristics and symptom profile of the Long COVID (LC) patient group.

### Association between PAF and pain severity in LC patients with new-onset chronic pain

A negative association was observed between global PAF (averaged across all electrode sites) and pain severity scores, but did not withstand adjustment for confounders (standardised β = −0.336, 95% CI [-0.88, 0.21], F(3,21) = 1.85, adj. R^2^ = 0.096, FDR-corrected p = 0.222 for max-PAF; standardised β = −0.327, 95% CI [-0.99, 0.34], F(3,21) = 1.88, adj. R^2^ = 0.099, FDR-corrected p = 0.229 for aperiodic-adjusted PAF) (Supplementary Materials Table S2).

Regional analysis indicated a significant relationship between pain severity and PAF over posterior electrode sites, even when adjusting for the potential confounding effects of age and depression (Table 2). The effect of introducing each confounding variable to the regression models is detailed in Supplementary Material Table S2. More specifically, lower posterior PAF estimates were associated with greater pain severity scores on the C19-YRS (standardised β = −0.602, 95% CI [−1.05, −0.15], coefficient p = 0.001 for max-PAF; standardised β = −0.581, 95% CI [−1.12, −0.04], coefficient p = 0.002 for aperiodic-adjusted PAF)(Figure 1). This finding was consistent across methods of estimation of PAF (F(3,21) = 5.94, adj. R^2^ = 0.382, FDR-corrected p = 0.017 for max-PAF; F(3,21) = 5.35, adj. R^2^ = 0.352,

**Figure 1.**
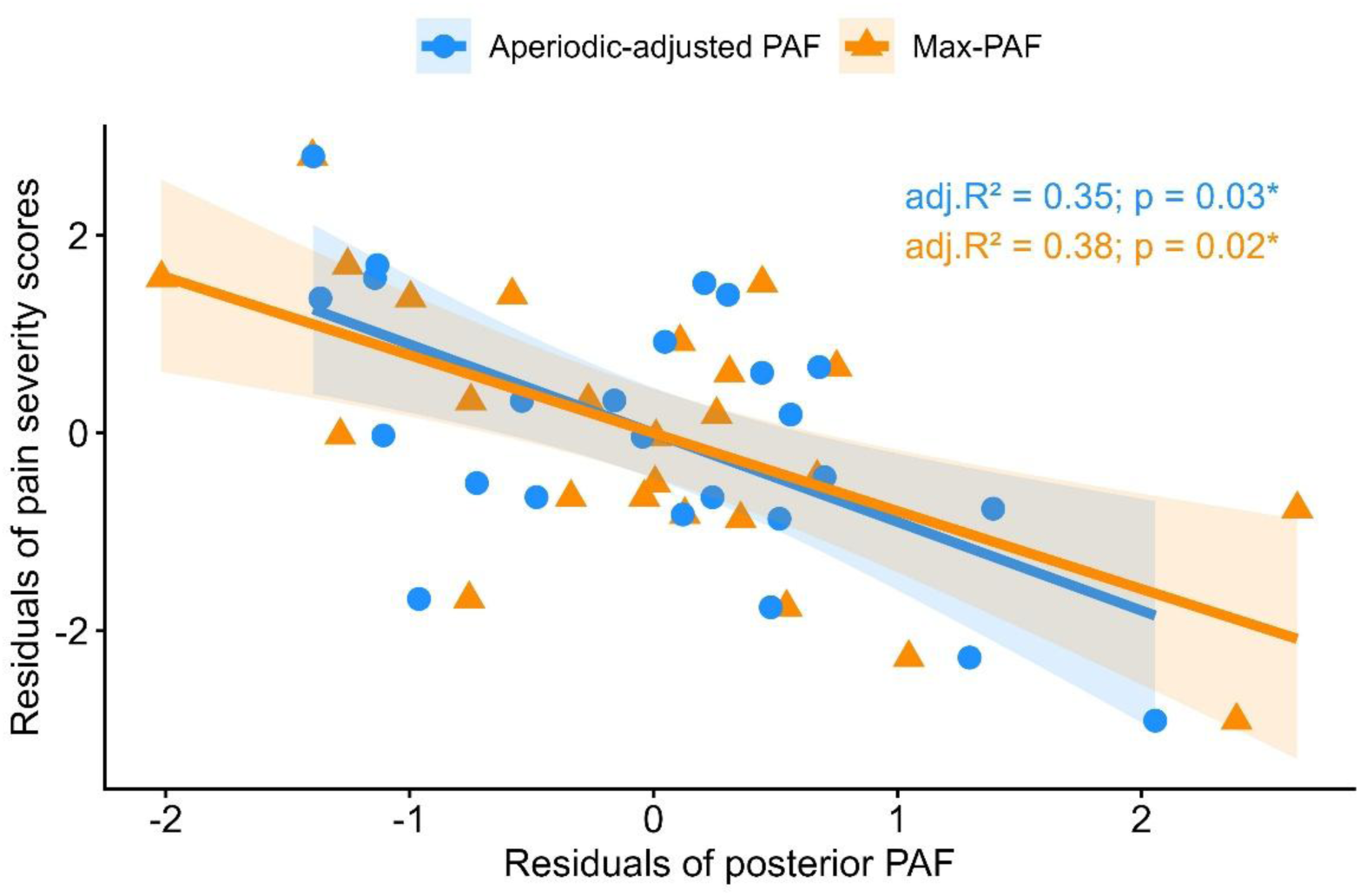
Partial regression plot showing association between peak alpha frequency (PAF) over posterior scalp region and pain severity in LC patients with new-onset chronic pain, controlling for the effects of age and depression. Each point represents a residual of C19-YRS pain severity scores (subscale of C19-YRS) after removing covariate effects, plotted against the residual of posterior PAF after removing the effects of the same covariates. Both max-PAF and aperiodic-adjusted PAF estimates are included in the plot.

FDR-corrected p = 0.027 for aperiodic-adjusted PAF). The independent metrics (PAF, age, depression) were not correlated between each other and in combination accounted for approximately 40% variance in pain intensity. No such association was observed for anterior (standardised β = −0.292, 95% CI [-0.86, 0.27], F(3,21) = 1.59, adj. R^2^ = 0.069, FDR-corrected p = 0.222 for max-PAF; standardised β = −0.163, 95% CI [-0.81, 0.48], F(3,21) = 1.15, adj. R^2^ = 0.018, FDR-corrected p = 0.352 for aperiodic-adjusted PAF) and central scalp regions (standardised β = −0.340, 95% CI [-0.89, 0.21], F(3,21) = 1.87, adj. R^2^ = 0.098, FDR-corrected p = 0.222 for max-PAF; standardised β = −0.326, 95% CI [-1.00, 0.35], F(3,21) = 1.83, adj. R^2^ = 0.094, FDR-corrected p = 0.229 for aperiodic-adjusted PAF).

As exploratory analysis, power spectral density of the strongest peak within the alpha range (PAF power spectral density) was also tested as a predictor, but it did not show a significant association with pain severity (standardised β = 0.049, 95% CI [-0.02, 0.12], F(3,21) = 0.94, adj. R^2^ = −0.008, p = 0.441 for max-PAF; standardised β = 0.009, 95% CI [-1.58, 1.60], F(3,21) = 0.92, adj. R^2^ = −0.011, p = 0.450 for aperiodic-adjusted PAF).

Due to consistency of findings across methods of PAF extraction and strong correlation between max-PAF and aperiodic-adjusted PAF estimates (r = 0.943, p < 0.001 for posterior

PAF; Figure 2) (see Supplementary Materials Table S3 for other regions), subsequent analyses results are reported here for aperiodic-adjusted PAF estimates only unless otherwise stated. Summary tables detailing results from both methods can be found in Supplementary Materials.

**Figure 2.**
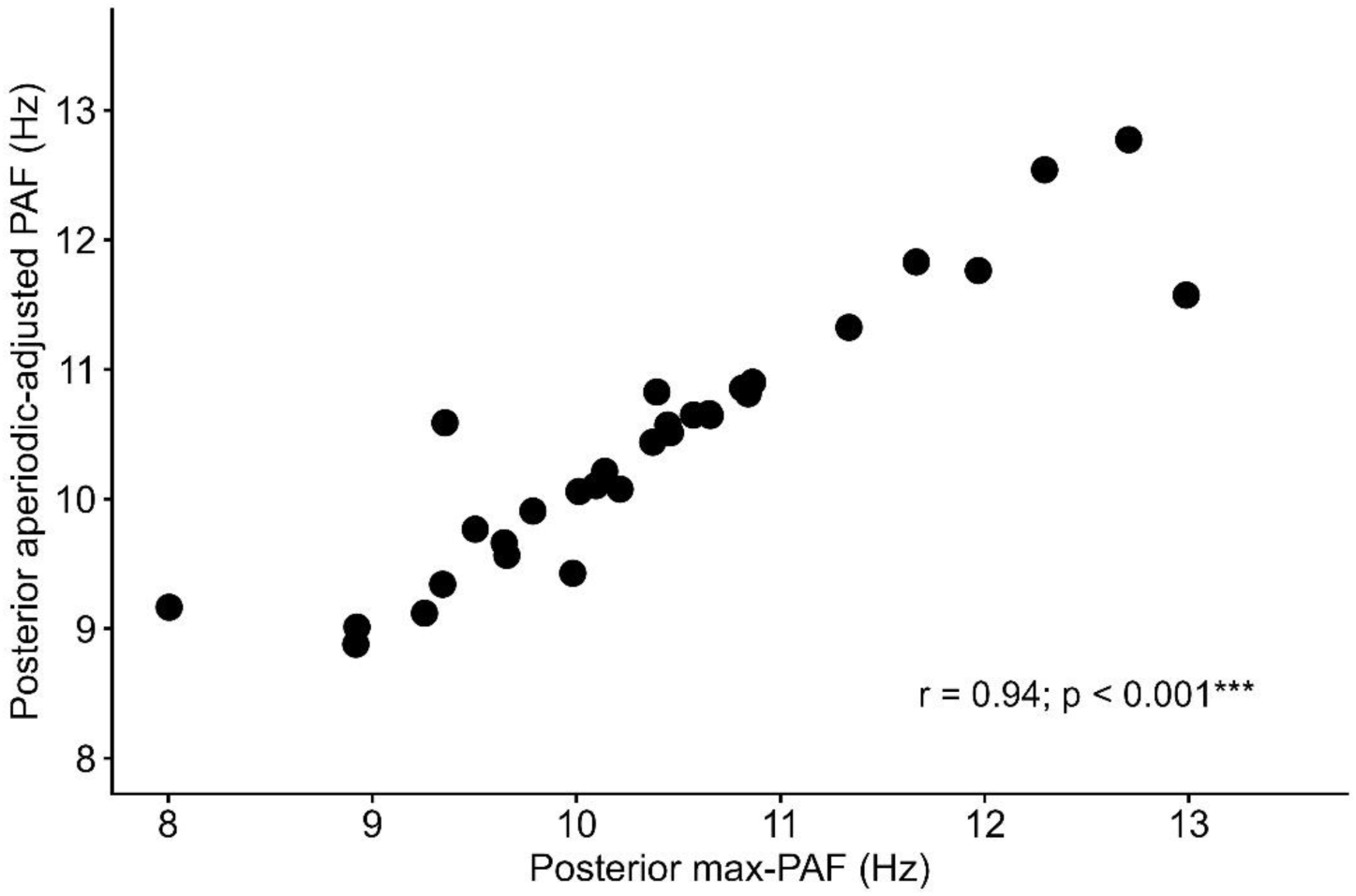
Scatterplot showing strong correlation between peak alpha frequency (PAF) estimates (posterior region) derived from two methods: frequency in the alpha range with maximum Welch’s spectral power (max-PAF) and frequency with the strongest Gaussian peak identified in the alpha range with spectral parameterisation algorithm (aperiodic-adjusted PAF). Correlation coefficient rho and p value based on Spearman’s rank correlation.

We tested whether PAF relates to other neurological symptoms frequently reported in LC and other pain-related metrics. Table 3 provides a summary of model outputs, with full model results presented in Supplementary Materials Table S4. No evidence was found for a relationship between posterior PAF and severity of fatigue (standardised β = −0.284, 95% CI [-0.92, 0.35], F(3,21) = 1.85, adj. R^2^ = 0.096, p = 0.168) or pain widespreadness (standardised β = −0.034, 95% [-1.15, 1.08], F(3,21) = 0.08, adj. R^2^ = −0.143, p = 0.968).

**Table 2.** Summary of multiple linear regression models on the association between posterior peak alpha frequency (PAF) and pain severity, controlling for the effects of age and depression. Models derived from max-PAF and aperiodic-adjusted PAF estimates are shown. Standardised β coefficients and standard errors for the independent variable and confounders, R^2^, adjusted R^2^, residual standard error (SE), F statistic and degrees of freedom, and false discovery rate (FDR)-corrected p values are reported for each model.

|  | Max-PAF | Aperiodic-adjusted PAF |
| --- | --- | --- |
| **Posterior PAF** | **-0.602\*\***<br>(0.216) | **-0.581\*\***<br>(0.262) |
| **Age** | -0.035<br>(0.021) | 0.059<br>(0.022) |
| **Depression** | 0.199<br>(0.044) | 0.268<br>(0.045) |
| **Constant** | 14.547***<br>(2.543) | 14.917***<br>(2.784) |
| **R<sup>2</sup>** | 0.459 | 0.433 |
| **Adj. R<sup>2</sup>** | 0.382 | 0.352 |
| **Residual SE** | 1.133 | 1.160 |
| **F Statistic** | 5.94 (df=3;21) | 5.35 (df=3;21) |
| **Corrected p** | **0.017\*** | **0.027\*** |
| *Note:* |  | *p**p***p<0.001 |

**Table 3.** Summary of multiple linear regression models on the association between posterior peak alpha frequency (PAF), other self-reported neurological symptoms of LC (fatigue and cognitive symptoms) and other pain-related metrics (pain duration and pain widespreadness). The models represented in the table are derived from aperiodic-adjusted PAF estimates. Standardised β coefficients and standard errors for the independent variable and confounders, R^2^, adjusted R^2^, residual standard error (SE), F statistic and degrees of freedom, and false discovery rate (FDR)-corrected p values are reported for each model.

|  | Severity of other neurological symptoms |  | Other pain-related metrics |  | Overall symptom severity |
| --- | --- | --- | --- | --- | --- |
|  | Fatigue | Cognition | Duration | Widespreadness |  |
| **Posterior PAF** | -0.284<br>(0.306) | -0.116<br>(0.572) | **-0.469\***<br>(41.772) | -0.034<br>(0.532) | -0.216<br>(2.572) |
| **Age** | -0.201<br>(0.026) | 0.043<br>(0.049) | 0.305<br>(3.612) | -0.019<br>(0.048) | -0.090<br>(0.221) |
| **Depression** | 0.233<br>(0.052) | **0.557\*\***<br>(0.098) | 0.209<br>(7.150) | -0.107<br>(0.096) | **0.735\*\*\***<br>(0.439) |
| **Constant** | 13.163***<br>(3.262) | 5.236<br>(6.091) | 1,159.825*<br>(453.172) | 7.966<br>(5.777) | 61.510*<br>(27.383) |
| **R<sup>2</sup>** | 0.209 | 0.340 | 0.330 | 0.013 | 0.630 |
| **Adj. R<sup>2</sup>** | 0.097 | 0.245 | 0.230 | -0.143 | 0.577 |
| **Residual Std. Error** | 1.359 | 2.537 | 183.680 | 2.341 | 11.405 |
| **F Statistic** | 1.85 (df=3;21) | 3.60 (df=3;21) | 3.29 (df=3;20) | 0.08 (df=3;19) | 11.91 (df=3;21) |
| **Model p value** | 0.168 | **0.030\*** | **0.042\*** | 0.968 | **<0.001\*\*\*** |
| Note: |  |  |  |  | *p**p***p<0.001 |

Furthermore, the results show that posterior PAF did not independently predict severity of cognitive impairments and overall LC symptom severity, with any models that reached significance threshold resulting from the confounding effects of depression (standardised β for PAF = −0.116, 95% CI [-1.31, 1.07], F(3,21) = 3.60, adj. R^2^ = 0.245, p = 0.030 for cognitive impairments; standardised β for PAF = −0.216, 95% CI [-5.57, 5.13], F(3,21) = 11.91, adj. R^2^ = 0.577, p < 0.001 for overall symptom severity)(Table 3). The data indicated a possible negative association between posterior PAF and duration of pain (standardised β = −0.469, 95% CI [-87.60, 86.67], F(3,21) = 3.29, adj. R^2^ = 0.230, p = 0.042 for aperiodic-adjusted PAF), however it was not sustained for max-PAF estimates after adjusting for confounders (standardised β = −0.387, 95% CI [-77.90, 77.13], F(3,21) = 2.38, adj. R^2^ = 0.153, p = 0.100 for max-PAF).

### Association between alpha band power and pain severity in LC patients with new-onset chronic pain

The results show no association between alpha band power and severity of pain symptoms at the global or regional level (Table 4). Detailed model outputs can be found in Supplementary Materials Table S5.

**Table 4.** Summary of linear regression models on the association between alpha band power and pain severity. Models derived from global and regional (including anterior, central and posterior regions) alpha band power estimates are shown. Standardised β coefficient and standard error for the independent variable, R^2^, adjusted R^2^, residual standard error, F statistic and degrees of freedom, and false discovery rate (FDR)-corrected p values are reported for each model.

|  | Global | Anterior | Central | Posterior |
| --- | --- | --- | --- | --- |
| **Alpha power spectral density (standardised)** | 0.063<br>(0.023) | 0.097<br>(0.022) | 0.010<br>(0.031) | -0.016<br>(0.009) |
| **Constant** | 6.822***<br>(0.440) | 6.785***<br>(0.411) | 6.904***<br>(0.463) | 6.940***<br>(0.394) |
| **$R^2$** | 0.004 | 0.009 | 0.000 | 0.000 |
| **Adj. $R^2$** | -0.039 | -0.034 | -0.043 | -0.043 |
| **Residual Std. Error** | 1.469 | 1.465 | 1.472 | 1.472 |
| **F Statistic** | 0.09 (df=1;23) | 0.22 (df=1;23) | 0.00 (df=1;23) | 0.01 (df=1;23) |
| **FDR-corrected p value** | 0.964 | 0.964 | 0.964 | 0.964 |
| *Note:* |  |  |  | * p ** p *** p<0.001 |

### Comparative analysis of PAF between LC patients with new-onset chronic pain and healthy pain-free controls

We next examined whether LC patients exhibited lower PAF compared to healthy pain-free controls across all predefined regions using multiple linear regression models, with a grouping factor and age as confounding variable. No evidence of significant differences was found between the overall LC patient group and the healthy pain-free control group for global PAF or PAF from any of the regions tested (Supplementary Materials Table S6). Patients were then grouped by severity of pain symptoms to enable severity-based subgroup analysis. The median cut-off of 7 was used to separate patients into a moderate pain subgroup consisting of patients reporting pain severity scores below median (n = 11, range of C19-YRS pain severity scores: 4–6, mean = 5.45, SD = 0.82), and a severe pain subgroup scoring equal or above median (n = 16, range of C19-YRS severity scores: 7–9, mean = 7.75, SD = 0.93). Results from the omnibus F-test on the 3-group (moderate pain subgroup, severe pain subgroup, control group) regression model indicated a significant effect of group on global PAF (F(2,54) = 3.39, p = 0.041) and regionally in the posterior scalp area (F(2,54) = 5.07, p = 0.010). Post-hoc pairwise comparison analysis revealed that patients reporting moderate pain presented significantly higher global and posterior PAF values compared to the healthy pain-free control group (for details of pairwise comparisons see Supplementary Materials Table S7). Moreover, additional differences localised to posterior scalp region were found between the pain severity subgroups, particularly significantly lower PAF values in the severe pain subgroup compared to the moderate pain subgroup (t(54) = −0.91, Tukey’s adjusted p = 0.021)(Figure 3). Our results show no significant differences between the severe pain subgroup and pain-free controls. Subgroup comparison of PAF estimates from the anterior region yielded inconsistent findings across PAF extraction methods. A significant main effect of group was found in association with max-PAF estimates from anterior scalp region (F(2,54) = 4.35, p = 0.018), with pairwise comparisons suggesting significantly increased anterior max-PAF in moderate pain subgroup compared to both severe pain subgroup and controls. Notwithstanding, this effect did not hold for aperiodic-adjusted PAF (F(2,54) = 3.05, p = 0.056). No differences were observed in the central region (for details of omnibus test see Supplementary Materials Table S6).

**Figure 3.**
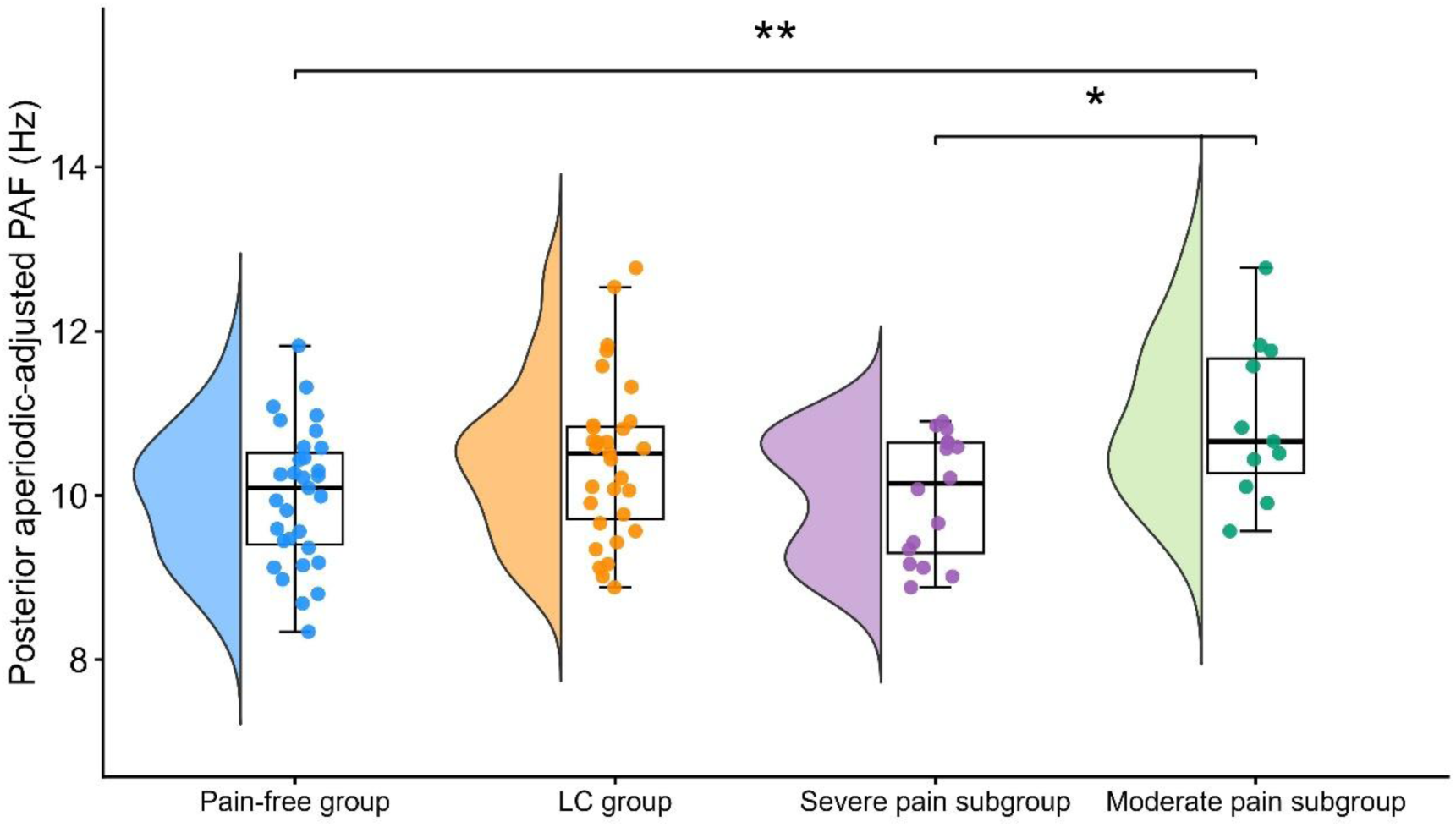
Comparison of peak alpha frequency (PAF) estimates over the posterior scalp region between LC patient group (n = 31) and healthy pain-free controls (n = 31). LC patient group is further stratified into subgroups based on median pain severity (median = 7) with a moderate pain subgroup including patients scoring below median value on the C19-YRS pain subscale (n = 11, range of pain severity scores: 4–6), and a severe pain subgroup including patients scoring equal or above median value (n = 16, range of pain severity scores: 7–9). Data is shown in raincloud plots combining un-mirrored violin plots, boxplots and individual data points to provide a comprehensive view of the distribution and spread of the data for each group and subgroup. Data represented in the figure is derived from aperiodic-adjusted PAF estimates. * p < 0.05; ** p < 0.01.

We also performed cluster-based permutation analysis on channel level data to confirm the findings from regional analysis. Comparison between the moderate pain subgroup and controls revealed one significant positive cluster covering occipital-parietal region and extending into left frontal and central electrode sites (p = 0.006, cluster statistic = 80.584) (Figure 4; bottom left panel). The results indicate electrodes sites where PAF was found to be significantly increased in patients with moderate pain as compared to pain-free controls. Additionally, the topographies differed between patient subgroups, revealing a significant decrease in PAF over occipital-parietal areas extending into central and left temporal electrode sites (p = 0.005, cluster statistic = −77.491) in patients with severe pain compared to the moderate pain subgroup (Figure 4.B; top right panel). No significant differences were observed between the severe pain patient subgroup and healthy controls (Figure 4.B; bottom right panel).

**Figure 4.**
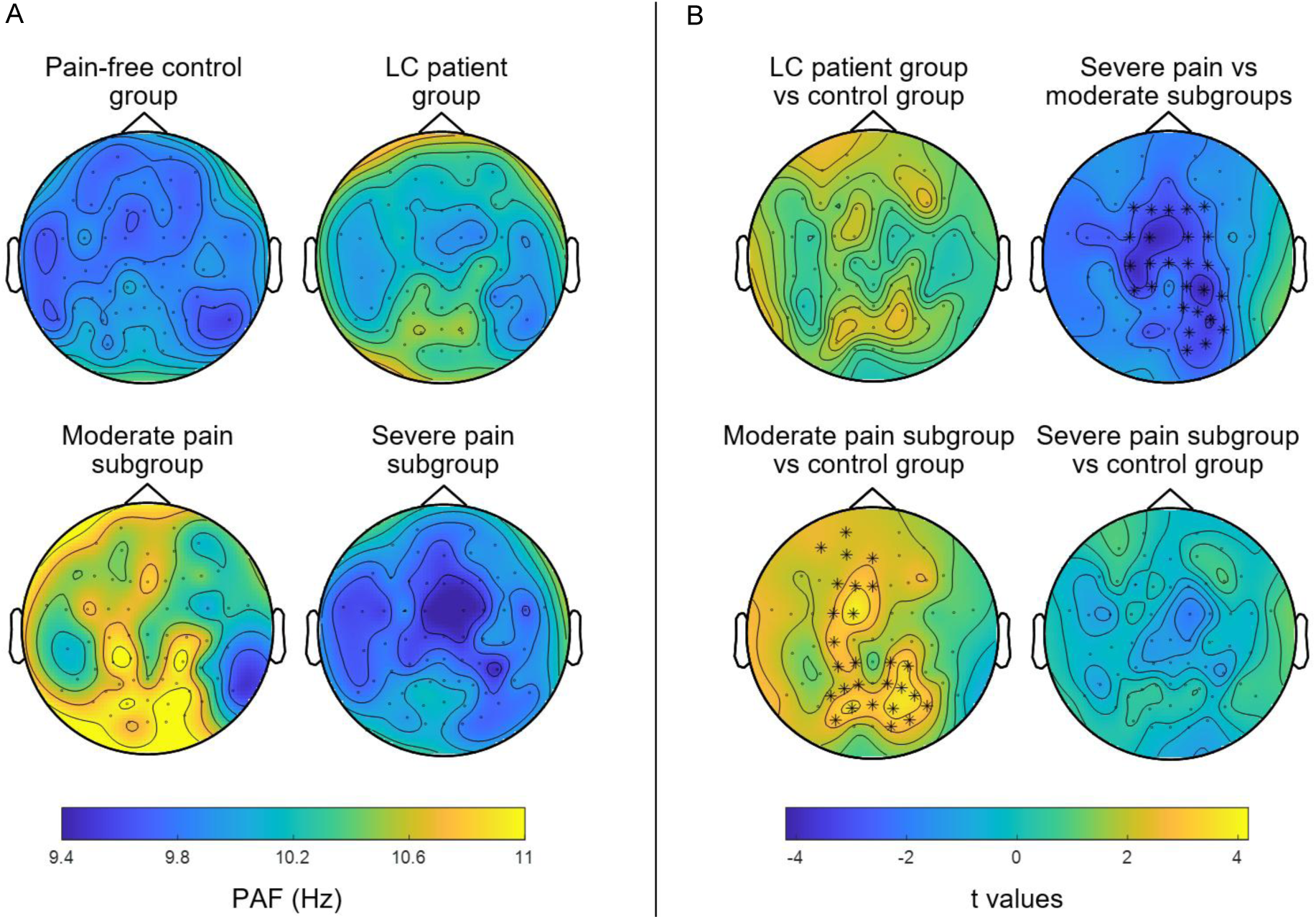
Cluster-based permutation comparison of peak alpha frequency (PAF) between groups at channel level. A) Topographical plots of PAF extracted for each electrode and averaged across participants within the healthy pain-free control group (n = 31), Long COVID (LC) patient group (n = 31), LC patient moderate pain subgroup (n = 11), and LC patient severe pain subgroup (n = 16). B) Channel-level comparison of PAF estimates between groups. Scalp maps represent topographical distribution of t-values for the comparison between groups, with warmer and colder colours representing increased and decreased PAF, respectively. Asterisks (*) denote electrode sites identified as part of significant clusters at significance level p < 0.05. Top left plot shows comparison between LC patient group and control group, with no significant clusters found. Top right plot shows comparison between severe pain and moderate pain LC patient subgroups, highlighting one significant negative cluster concentrated over central to centro-parietal areas and extending into frontal and right-lateralised occipital-parietal electrode sites. Bottom left panel shows comparison between patients reporting moderate pain and controls, highlighting one significant positive cluster covering occipital-parietal region and extending into left frontal and central electrode sites. Bottom right panel shows comparison between patients reporting severe pain and controls, with no significant clusters found. Data represented in the figure is derived from aperiodic-adjusted PAF estimates.

### Comparative analysis of alpha band power between LC patients with new-onset chronic pain and healthy pain-free controls

Alpha band power did not differ between the LC patient group and the control group at any of the regions tested, nor were any significant differences found between moderate and severe pain subgroups (see Supplementary Materials Table S8 for detailed omnibus test results).

## Discussion

To our knowledge, this is the first study to investigate EEG alpha oscillatory features associated with pain symptomology in a cohort of LC patients with new-onset chronic pain.

Consistent with our hypothesis, we found that lower PAF is associated with higher severity of LC-chronic pain. This result was localised to the posterior scalp region and consistent across different PAF estimation methods. Subgroup analysis revealed that LC patients with moderate pain presented significantly higher posterior PAF compared to both the severe pain subgroup and healthy pain-free controls. Our results on alpha band power show no association with pain severity, as well as no differences between groups.

### Association between new-onset LC-chronic pain and EEG alpha oscillatory features

While studies have shown decreased PAF in chronic pain patients, evidence of an inverse relation between PAF and pain severity comes mostly from human experimental pain studies [15,20,21]. Our findings extend the literature on the PAF-pain association to a novel post-infection pain syndrome. Here we report that lower PAF in the posterior scalp region is associated with higher severity of LC-chronic pain, independent of age and depression-related effects on PAF. Alpha oscillations are thought to play a role in the dynamic gating and processing of sensory information by functionally inhibiting task- or stimulus-irrelevant cortical regions [13]. Thus, slower alpha activity may reflect impaired ability to modulate incoming pain information contributing to heightened pain severity [20,21]. The regional specificity of our finding could be explained by alpha rhythms being most prominent over occipital-parietal scalp region, supporting the idea that posterior scalp sites are where alpha/PAF differences and behaviour associations are most evident [43]. This is corroborated by previous research showing differences in PAF between chronic pain patients and pain-free controls are more pronounced in the occipital-parietal areas [42,65].

Max-PAF and aperiodic-adjusted PAF estimates were highly correlated, as previously shown by Deodato *et al.* [12], and yielded comparable results. This demonstrates the robustness of our findings across methods and indicates that the observed association is not attributable to the impact of aperiodic 1/f-like activity on PAF estimation. Our results further complement prior studies on PAF-pain association, which have not accounted for the effects of the aperiodic component in their analysis [15,20,21,44].

Furthermore, we show that posterior PAF is not associated with severity of fatigue, cognitive impairments, and overall LC symptom severity, providing evidence to support symptom-specificity of our findings. This builds upon the framework outlined in our prior review of quantitative EEG studies on LC and similar multisymptom clinical syndromes [59], where we propose a symptom-focused approach to LC research. The observed negative association between pain duration and posterior PAF aligns with the observations by De Vries *et al.* [65]. Nevertheless, this exploratory finding is unlikely to withstand correction for multiple testing and needs confirming in future longitudinal studies.

### Comparison between LC patients with new-onset chronic pain and pain-free controls

To date, no study has investigated the association between LC pain and EEG oscillatory activity. Limited prior work has investigated alpha oscillatory cortical neural activity in LC (without emphasis on pain). These studies focused mainly on post-COVID cognitive disturbances and reported mixed results of reduced [1], increased [37] or no difference [8,53] in alpha power/current source density in patients compared to controls or EEG recordings from the same individuals recorded prior to developing LC. Only a single study has incorporated PAF in the analysis, with findings suggesting reduced occipital PAF in patients with post-COVID cognitive disturbances compared to controls [8].

Our study found no differences in PAF or alpha band power between the overall LC patient group and controls. Several possible explanations for the observed lack of differences could be proposed: (1) there are effectively no differences in EEG alpha oscillatory features between groups; (2) there are subtle differences between groups, although the small sample size limits the ability to detect them; and (3) heterogeneity within the patient group could mask potential neurophysiological differences that are present only in a subset of patients.

An interesting finding of our study is that LC patients with moderate pain present heightened PAF on average compared to pain-free controls. Notably, we also found that LC patients with severe pain have significantly reduced posterior PAF compared to moderate pain patients, but not compared to controls as hypothesised. An interpretation of these findings is that individuals with slower PAF are more vulnerable or predisposed to develop severe pain symptoms upon contraction of COVID-19 infection. Accordingly, the significantly faster PAF observed in LC patients with moderate pain may have served as a protective mechanism against heightened pain sensitivity. This is supported by evidence that resting-state pain-free PAF has predictive value on the severity of prolonged pain in experimental tonic pain models and clinical post-operative pain [20,21,44]. Validation of this hypothesis would require measurement of pre-COVID PAF from EEG signals collected prior to development of LC symptoms, though such data are not available for this cohort. An alternative explanation is that subgroup differences in PAF may indicate state-dependent neurophysiological changes that reflect fluctuations in pain over time. Though plausible in view of the evidence on PAF-pain association, studies have failed to demonstrate that PAF tracks changes in pain over time [20,29] reinforcing prior reports of PAF as a relatively stable neurophysiological trait marker [2,25].

Additionally, we have previously reported that 85% of patients from our LC cohort show signs of central sensitisation based on quantitative sensory testing (QST) profiling, supporting the presence of nociplastic type pain [36]. It is possible that the remaining proportion of patients present with pain associated with different mechanistic features, with likely dominance of nociceptive and neuropathic contributors which can be associated with different neurophysiological patterns. Further to this, heterogeneity could also be attributed, at least in part, to the presence of distinct LC symptom phenotypes. This possibility merits further investigation in phenotype-specific cohorts, either in separate studies or combined for comparative analysis.

## Limitations

This study has several limitations that should be considered when interpreting the findings. First, the small sample size may limit generalization of findings. We specifically recruited LC patients without pre-existing musculoskeletal chronic pain prior to COVID-19 infection in an effort to prevent confounding effects of pain of different aetiology, which introduced an additional challenge to recruitment process and limited the pool of potential participants.

Second, subgrouping patients based on pain severity may reduce statistical power and introduce classification bias. On this note, our median cut-off (median = 7) between moderate and severe pain subgroups is consistent with that suggested in chronic pain literature [4,7,31,57] and often used in clinic, thus supporting the clinical relevance of our findings. Nevertheless, the results from subgroup analysis should be regarded as exploratory and preliminary, warranting further validation in larger cohorts. Third, eyes-open resting-state EEG was selected to reduce drowsiness and maintain attention, which is particularly important in the LC population where fatigue and sleep disturbances may influence alertness. While this approach is methodologically valid, it should be noted that alpha power is typically higher in the eyes-closed condition and PAF shifts can occur, limiting direct comparison with eyes-closed resting-state EEG studies. Finally, we acknowledge the potential for confounding by other factors not measured or modelled in this study, such as sleep, anxiety and comorbidities.

### Conclusions and recommendations for future research

The findings of this study provide evidence to support posterior PAF as an objective, measurable neurophysiological brain feature that associates with severity of new-onset LC-chronic pain. This work contributes to the growing body of evidence suggesting, among other factors, a neurophysiological basis of chronic pain symptoms in LC, a condition that to date remains the subject of stigma and associations with psychosomatisation. Larger, longitudinal design studies are needed to assess how PAF dynamically evolves alongside the natural progression of pain and to further elucidate subgroup differences in PAF. Overall, our findings, while early, suggest non-invasive targeted neuromodulation could be explored as a potential therapeutic avenue for new-onset LC chronic pain. Tailored neuromodulation strategies show promise within an integrated multidisciplinary management framework for LC-chronic pain, and could be relevant to other similar post-infectious syndromes manifesting with chronic pain symptomology.

## Data availability statement

All data are available upon reasonable request to the corresponding author.

## Author contributions

OK and MS designed the MUSLOC study, and BSP contributed to design of the EEG component of the study. BSP and OK collected the data. BSP, ID, CB and AJC were involved in planning analytical methodology. BSP cleaned and analysed the data. BSP drafted the manuscript. All authors critically revised the manuscript and approved the version for publication. MS provided clinical input as a clinician managing patients with the condition in his practice.

## Data Availability

All data are available upon reasonable request to the corresponding author.

## Acknowledgements

The authors would like to express their heartfelt gratitude to all participants and their families for their invaluable contribution to this study. Special thanks to the staff of the LCH Long COVID service for their crucial help in recruiting participants.

