## Supplementary materials for "Peak alpha frequency is associated with pain severity in Long COVID patients with new-onset chronic pain"

Authors: $\text{Bárbara Silva-Passadouro}^{\text{1}}$, $\text{Omar Khoja}^{\text{1}},$ $\text{Alexander J. Casson}^{2}$, $\text{Ioannis Delis}^{\text{3,4}}$,$\text{ Christopher Brown}^{5}$, $\text{Manoj Sivan}^{\text{1,6}}$

1 Leeds Institute of Rheumatology and Musculoskeletal Medicine, School of Medicine, University of Leeds, Leeds, UK
2 Department of Electrical and Electronic Engineering, University of Manchester, Manchester, UK
3 School of Biomedical Sciences, Faculty of Biological Sciences, University of Leeds, Leeds, UK
4 School of Electrical and Computer Engineering, National Technical University of Athens, Athens, Greece
5 Department of Psychology, Institute of Population Health, University of Liverpool, Liverpool, UK
6 National Demonstration Centre in Rehabilitation Medicine, Leeds Teaching Hospitals NHS Trust, Leeds, UK

**Table S1. Summary of pre- and post-COVID symptom severity in the Long COVID patient group based C19-YRS items.**

| **Long COVID symptoms** |  |  |
| --- | --- | --- |
| C19-YRS items (missing: 4) | Pre-COVID | Post-COVID |
| Breathlessness - at rest | 4 participants (14.8%) | 17 participants (63.0%) |
| mean severity (SD) | 4.2 (3.3) | 4.6 (2.8) |
| Breathlessness - dressing | 6 participants (22.2%) | 23 participants (85.2%) |
| mean severity (SD) | 2.8 (1.8) | 4.9 (2.9) |
| Breathlessness - stairs | 13 participants (48.1%) | 26 participants (96.3%) |
| mean severity (SD) | 1.9 (0.9) | 6.2 (2.6) |
| Cough/throat sensitivity/voice change | 5 participants (18.5%) | 19 participants (70.4%) |
| mean severity (SD) | 3.2 (2.6) | 5.4 (2.6) |
| Fatigue | 16 participants (59.3%) | 27 participants (100%) |
| mean severity (SD) | 2.1 (0.8) | 8.2 (1.4) |
| Continence | 5 participants (18.5%) | 13 participants (48.1%) |
| mean severity (SD) | 2.0 (1.7) | 5.6 (2.2) |
| Pain | 0 participants (0%) | 27 participants (100%) |
| mean severity (SD) | 0.0 (0.0) | 6.8 (1.4) |
| Cognition | 12 participants (44.4%) | 25 participants (92.6%) |
| mean severity (SD) | 2.5 (1.4) | 6.4 (2.3) |
| Anxiety | 16 participants (59.3%) | 25 participants (92.6%) |
| mean severity (SD) | 2.7 (1.6) | 6.1 (2.5) |
| Depression | 9 participants (33.3%) | 19 participants (70.4%) |
| mean severity (SD) | 3.4 (2.3) | 6.0 (2.1) |
| PTSD | 5 participants (18.5%) | 16 participants (59.3%) |
| mean severity (SD) | 1.8 (1.8) | 5.4 (2.5) |
| Communication | 6 participants (22.2%) | 20 participants (74.1%) |
| mean severity (SD) | 1.7 (0.8) | 5.6 (2.6) |
| Mobility | 5 participants (18.5%) | 21 participants (77.8%) |
| mean severity (SD) | 1.8 (1.8) | 5.9 (2.3) |
| Personal care | 2 participants (7.4%) | 14 participants (51.8%) |
| mean severity (SD) | 3.5 (3.5) | 3.9 (2.4) |
| Other activities of daily living | 5 participants (18.5%) | 25 participants (92.6%) |
| mean severity (SD) | 1.6 (0.9) | 7.1 (1.8) |
| Social role | 2 participants (7.4%) | 25 participants (92.6%) |
| mean severity (SD) | 1.0 (0.0) | 5.9 (2.3) |
| C19-YRS symptom severity, mean (SD) | 7.2 (8.2) | 45.1 (17.4) |
| C19-YRS functional disability, mean (SD) | 1.3 (2.9) | 22.8 (10.9) |
| C19-YRS global health score, mean (SD) | 8.0 (1.2) | 2.7 (1.6) |

**Table S2. Summary of multiple linear regression models on the association between peak alpha frequency (PAF) and pain severity.** Model 1 represents results of simple linear regression between dependent variable (pain severity) and independent variable (PAF). Models 2 and 3 outline the effects of introducing each confounding variable (age and depression, respectively); and Model 4 includes confounding effects of both age and depression. Models derived from max-PAF and aperiodic-adjusted PAF estimates are shown. Standardised β coefficients and 95% confidence intervals (CI) for each independent variable, F statistic and degrees of freedom, adjusted $\text{R}^{\text{2}}$, raw overall model p values and false discovery rate (FDR)-corrected p values are reported for each combination of PAF extraction method (max-PAF and aperiodic-adjusted PAF), scalp region (global, anterior, central and posterior region), and model (model 1 to model 4). Only the p values from the models adjusted for both confounders (model 4) were adjusted for multiple testing using FDR correction. * p < 0.05; ** p < 0.01; *** p < 0.001.

|  | **Standardised β coefficient for PAF [95% CI]** | **Standardised β coefficient for age [95% CI]** | **Standardised β coefficient for depression**  **[95% CI]** | **F statistic (degrees of freedom)** | **Adj.** $\text{R}^{\text{2}}$ | **Overall model p value** | **FDR-corrected p value** |
| --- | --- | --- | --- | --- | --- | --- | --- |
| **Global** | | | | | | | |
| **max-PAF models** | | | | | | | |
| Model 1 | **-0.402*** [-0.89 0.09] |  |  | 4.42 (1,23) | 0.125 | **0.047*** | --- |
| Model 2 | -0.417 [-0.92 0.08] | -0.111 [-0.16 -0.06] |  | 2.31 (2,22) | 0.098 | 0.123 | --- |
| Model 3 | -0.320 [-0.85 0.21] |  | 0.206 [0.09 0.32] | 2.70 (2,22) | 0.124 | 0.089 | --- |
| Model 4 | -0.336 [-0.88 0.21] | -0.111 [-0.17 -0.06] | 0.206 [0.09 0.32] | 1.85 (3,21) | 0.096 | 0.168 | 0.222 |
| **aperiodic-adjusted PAF models** | | | | | | | |
| Model 1 | **-0.401*** [-1.02 0.22] |  |  | 4.42 (1,23) | 0.125 | **0.047*** | --- |
| Model 2 | -0.401 [-1.03 0.23] | -0.050 [-0.10 0.01] |  | 2.15 (2,22) | 0.088 | 0.140 | --- |
| Model 3 | -0.329 [-0.98 0.32] |  | 0.228 [0.12 0.34] | 2.88 (2,22) | 0.136 | 0.077 | --- |
| Model 4 | -0.327 [-0.99 0.34] | -0.063 [-0.12 -0.01] | 0.231 [0.12 0.35] | 1.88 (3,21) | 0.099 | 0.164 | 0.229 |
| **Anterior region** | | | | | | | |
| **max-PAF models** | | | | | | | |
| Model 1 | -0.375 [-0.87 0.12] |  |  | 3.77 (1,23) | 0.103 | 0.064 | --- |
| Model 2 | -0.382 [-0.89 0.13] | -0.084 [-0.14 -0.03] |  | 1.91 (2,22) | 0.070 | 0.172 | --- |
| Model 3 | -0.286 [-0.84 0.27] |  | 0.211 [0.09 0.33] | 2.37 (2,22) | 0.103 | 0.117 | --- |
| Model 4 | -0.292 [-0.86 0.27] | -0.088 [-0.14 -0.03] | 0.213 [0.09 0.33] | 1.59 (3,21) | 0.069 | 0.222 | 0.222 |
| **aperiodic-adjusted PAF models** | | | | | | | |
| Model 1 | -0.231 [-0.86 0.40] |  |  | 1.29 (1,23) | 0.012 | 0.267 | --- |
| Model 2 | -0.228 [-0.87 0.42] | -0.037 [-0.09 0.02] |  | 0.64 (2,22) | -0.031 | 0.539 | --- |
| Model 3 | -0.168 [-0.80 0.46] |  | 0.297 [0.18 0.41] | 1.76 (2,22) | 0.059 | 0.196 | --- |
| Model 4 | -0.163 [-0.81 0.48] | -0.057 [-0.11 -0.01] | 0.301 [0.18 0.42] | 1.15 (3,21) | 0.018 | 0.352 | 0.352 |
| **Central region** | | | | | | | |
| **max-PAF models** | | | | | | | |
| Model 1 | **-0.408*** [-0.90 0.08] |  |  | 4.60 (1,23) | 0.131 | **0.043*** | --- |
| Model 2 | **-0.421*** [-0.92 0.08] | -0.104 [-0.16 -0.05] |  | 2.37 (2,22) | 0.103 | 0.116 | --- |
| Model 3 | -0.327 [-0.86 0.21] |  | 0.199 [0.08 0.32] | 2.75 (2,22) | 0.127 | 0.086 | --- |
| Model 4 | -0.340 [-0.89 0.21] | -0.105 [-0.16 -0.05] | 0.200 [0.08 0.32] | 1.87 (3,21) | 0.098 | 0.165 | 0.222 |
| **aperiodic-adjusted PAF models** | | | | | | | |
| Model 1 | **-0.404*** [-1.02 0.21] |  |  | 4.50 (1,23) | 0.127 | **0.045*** | --- |
| Model 2 | **-0.405*** [-1.03 0.22] | -0.058 [-0.11 -0.01] |  | 2.20 (2,22) | 0.091 | 0.134 | --- |
| Model 3 | -0.326 [-0.98 0.33] |  | 0.213 [0.10 0.33] | 2.80 (2,22) | 0.131 | 0.082 | --- |
| Model 4 | -0.326 [-1.00 0.35] | -0.070 [-0.12 -0.01] | 0.217 [0.10 0.33] | 1.83 (3,21) | 0.094 | 0.172 | 0.229 |
| **Posterior region** | | | | | | | |
| **max-PAF models** | | | | | | | |
| Model 1 | **-0.649***** [-1.08 -0.22] |  |  | 16.72 (1,23) | 0.396 | **<0.001***** | --- |
| Model 2 | **-0.648***** [-1.09 -0.21] | -0.022 [-0.07 0.02] |  | 8.01 (2,22) | 0.369 | **0.002**** | --- |
| Model 3 | **-0.605**** [-1.04 -0.17] |  | 0.197 [0.11 0.29] | 9.28 (2,22) | 0.408 | **0.001**** | --- |
| Model 4 | **-0.602**** [-1.05 -0.15] | -0.035 [-0.08 0.01] | 0.199 [0.11 0.29] | 5.94 (3,21) | 0.382 | **0.004**** | **0.017*** |
| **aperiodic-adjusted PAF models** | | | | | | | |
| Model 1 | **-0.597**** [-1.13 -0.06] |  |  | 12.74 (1,23) | 0.328 | **0.002**** | --- |
| Model 2 | **-0.614**** [-1.17 -0.06] | 0.080 [0.03 0.13] |  | 6.26 (2,22) | 0.305 | **0.007**** | --- |
| Model 3 | **-0.568**** [-1.09 -0.05] |  | 0.273 [0.18 0.36] | 8.30 (2,22) | 0.378 | **0.002**** | --- |
| Model 4 | **-0.581**** [-1.12 -0.04] | 0.059 [0.01 0.10] | 0.268 [0.17 0.36] | 5.35 (3,21) | 0.352 | **0.007**** | **0.027*** |

**Table S3. Correlation between max-PAF estimates and aperiodic-adjusted PAF estimates derived from global, anterior, central and posterior regions.** *** p < 0.001.

| **Region** | **Spearman’s rho** | **p value** |
| --- | --- | --- |
| Global | 0.960 | **<0.001***** |
| Anterior | 0.925 | **<0.001***** |
| Central | 0.975 | **<0.001***** |
| Posterior | 0.943 | **<0.001***** |

**Table S4. Summary of multiple linear regression models on the association between posterior peak alpha frequency (PAF), other self-reported neurological symptoms of Long COVID, pain-related metrics and overall symptom severity.** Model 1 represents results of simple linear regression between dependent variable (severity of fatigue, severity of cognitive symptoms, pain duration, pain widespreadness, overall LC symptom severity) and independent variable (PAF over posterior scalp region). Models 2 and 3 outline the effects of introducing each confounding variable (age and depression, respectively); and Model 4 includes confounding effects of both age and depression. Models derived from max-PAF and aperiodic-adjusted PAF estimates are shown. Standardised β coefficients and 95% confidence intervals (CI) for each independent variable, F statistic and degrees of freedom, adjusted $\text{R}^{\text{2}}$ and raw overall model p values are reported for each combination of dependent variable tested (severity of fatigue, severity of cognitive symptoms, pain duration, pain widespreadness or overall LC symptom severity), PAF extraction method (max-PAF and aperiodic-adjusted PAF), and model (model 1 to model 4). No FDR correction for multiple testing was applied to these models due to exploratory nature of this analysis. * p < 0.05; ** p < 0.01; *** p < 0.001.

|  | **Standardised β coefficient for posterior PAF**  **[95% CI]** | **Standardised β coefficient for age [95% CI]** | **Standardised β coefficient for depression**  **[95% CI]** | **F statistic (degrees of freedom)** | **Adj.** $\text{R}^{\text{2}}$ | **Overall model p value** |
| --- | --- | --- | --- | --- | --- | --- |
| **Posterior max-PAF** | | | | | | |
| **Fatigue** | | | | | | |
| Model 1 | -0.325 [-0.85 0.20] |  |  | 2.72 (1,23) | 0.067 | 0.113 |
| Model 2 | -0.314 [-0.84 0.21] | -0.235 [-0.29 -0.18] |  | 2.11 (2,22) | 0.085 | 0.145 |
| Model 3 | -0.282 [-0.83 0.26] |  | 0.189 [0.08 0.30] | 1.78 (2,22) | 0.061 | 0.191 |
| Model 4 | -0.267 [-0.81 0.27] | -0.248 [-0.30 -0.19] | 0.206 [0.09 0.32] | 1.76 (3,21) | 0.087 | 0.186 |
| **Cognitive impairments** | | | | | | |
| Model 1 | -0.191 [-1.31 0.93] |  |  | 0.87 (1,23) | -0.005 | 0.360 |
| Model 2 | -0.194 [-1.34 0.96] | 0.056 [-0.06 0.17] |  | 0.45 (2,22) | -0.048 | 0.641 |
| Model 3 | -0.066 [-1.05 0.92] |  | **0.557**** [0.35 0.76] | 5.43 (2,22) | 0.270 | **0.012*** |
| Model 4 | -0.067 [-1.08 0.94] | 0.021 [-0.08 0.12] | **0.555**** [0.35 0.76] | 3.46 (3,21) | 0.235 | **0.035*** |
| **Pain duration** | | | | | | |
| Model 1 | **-0.425*** [-75.45 74.60] |  |  | 4.86 (1,23) | 0.144 | **0.038*** |
| Model 2 | **-0.431*** [-74.92 74.06] | 0.230 [-7.46 7.92] |  | 3.21 (2,22) | 0.161 | 0.061 |
| Model 3 | -0.380 [-78.31 77.55] |  | 0.183 [-15.90 16.27] | 2.83 (2,22) | 0.137 | 0.081 |
| Model 4 | -0.387 [-77.90 77.13] | 0.225 [-7.53 7.98] | 0.176 [-15.82 16.17] | 2.38 (3,21) | 0.153 | 0.100 |
| **Pain widespreadness** | | | | | | |
| Model 1 | 0.103 [-0.77 0.97] |  |  | 0.23 (1,23) | -0.036 | 0.639 |
| Model 2 | 0.103 [-0.79 1.00] | -0.038 [-0.13 0.06] |  | 0.12 (2,22) | -0.087 | 0.885 |
| Model 3 | 0.084 [-0.83 1.00] | -0.087 [-0.28 0.11] |  | 0.18 (2,22) | -0.080 | 0.835 |
| Model 4 | 0.084 [-0.86 1.03] | -0.028 [-0.13 0.07] | -0.083 [-0.29 0.12] | 0.12 (3,21) | -0.136 | 0.947 |
| **Overall symptom severity** | | | | | | |
| Model 1 | -0.311 [-6.84 6.22] |  |  | 2.46 (1,23) | 0.057 | 0.131 |
| Model 2 | -0.307 [-6.98 6.37] | -0.083 [-0.76 0.60] |  | 1.27 (2,22) | 0.022 | 0.301 |
| Model 3 | -0.149 [-4.79 4.49] |  | **0.719***** [-0.23 1.67] | 15.69 (2,22) | 0.550 | **<0.001***** |
| Model 4 | -0.141 [-4.81 4.53] | -0.129 [-0.59 0.33] | **0.728***** [-0.23 1.69] | 10.7 (3,21) | 0.548 | **<0.001***** |
| **Posterior aperiodic-adjusted PAF** | | | | | | |
| **Fatigue** | | | | | | |
| Model 1 | -0.352 [-0.97 0.26] |  |  | 3.26 (1,23) | 0.086 | 0.084 |
| Model 2 | -0.313 [-0.95 0.32] | -0.183 [-0.24 -0.13] |  | 2.03 (2,22) | 0.079 | 0.155 |
| Model 3 | -0.329 [-0.95 0.29] |  | 0.218 [0.11 0.33] | 2.27 (2,22) | 0.096 | 0.127 |
| Model 4 | -0.284 [-0.92 0.35] | -0.201 [-0.26 -0.15] | 0.233 [0.12 0.34] | 1.85 (3,21) | 0.096 | 0.168 |
| **Cognitive impairments** | | | | | | |
| Model 1 | -0.166 [-1.50 1.16] |  |  | 0.65 (1,23) | -0.015 | 0.428 |
| Model 2 | -0.185 [-1.58 1.21] | 0.087 [-0.03 0.21] |  | 0.40 (2,22) | -0.053 | 0.678 |
| Model 3 | -0.107 [-1.24 1.02] |  | **0.560**** [0.36 0.76] | 5.61 (2,22) | 0.278 | **0.011*** |
| Model 4 | -0.116 [-1.31 1.07] | 0.043 [-0.06 0.14] | **0.557**** [0.35 0.76] | 3.60 (3,21) | 0.245 | **0.030*** |
| **Pain duration** | | | | | | |
| Model 1 | **-0.438*** [-88.66 87.79] |  |  | 5.21 (1,23) | 0.155 | **0.032*** |
| Model 2 | **-0.498*** [-87.11 86.12] | 0.316 [-7.24 7.87] |  | 4.24 (2,22) | 0.220 | **0.028*** |
| Model 3 | **-0.408*** [-88.91 88.09] |  | 0.224 [-15.21 15.66] | 3.33 (2,22) | 0.169 | 0.055 |
| Model 4 | **-0.469*** [-87.60 86.67] | 0.305 [-7.23 7.84] | 0.209 [-14.71 15.12] | 3.29 (3,21) | 0.230 | **0.042*** |
| **Pain widespreadness** | | | | | | |
| Model 1 | -0.026 [-1.06 1.01] |  |  | 0.01 (1,23) | -0.047 | 0.905 |
| Model 2 | -0.020 [-1.10 1.06] | -0.035 [-0.13 0.06] |  | 0.02 (2,22) | -0.098 | 0.982 |
| Model 3 | -0.038 [-1.10 1.03] |  | -0.110 [-0.30 0.08] | 0.13 (2,22) | -0.086 | 0.881 |
| Model 4 | -0.034 [-1.15 1.08] | -0.019 [-0.12 0.08] | -0.107 [-0.31 0.09] | 0.08 (3,21) | -0.143 | 0.968 |
| **Overall symptom severity** | | | | | | |
| Model 1 | -0.313 [-8.01 7.38] |  |  | 2.50 (1,23) | 0.059 | 0.127 |
| Model 2 | -0.306 [-8.38 7.77] | -0.032 [-0.73 0.66] |  | 1.21 (2,22) | 0.017 | 0.317 |
| Model 3 | -0.236 [-5.37 4.90] |  | **0.728***** [-0.17 1.62] | 18.11 (2,22) | 0.588 | **<0.001***** |
| Model 4 | -0.216 [-5.57 5.13] | -0.090 [-0.55 0.37] | **0.735***** [-0.18 1.65] | 11.91 (3,21) | 0.577 | **<0.001***** |

**Table S5. Summary of multiple linear regression models on the association between** **alpha band power and pain severity.** Model 1 represents results of simple linear regression between dependent variable (pain severity) and independent variable (alpha band power). Models 2 and 3 outline the effects of introducing each confounding variable (age and depression, respectively); and Model 4 includes confounding effects of both age and depression. Standardised β coefficients and 95% confidence intervals (CI) for each independent variable, F statistic and degrees of freedom, adjusted $\text{R}^{\text{2}}$ and raw overall model p values are reported for each scalp region (global, anterior, central and posterior region), and model (model 1 to model 4). Only the p values from the models adjusted for both confounders (model 4) were adjusted for multiple testing using FDR correction.

|  | **Standardised β coefficient for alpha band power [95% CI]** | **Standardised β coefficient for age [95% CI]** | **Standardised β coefficient for depression**  **[95% CI]** | **F statistic (degrees of freedom)** | **Adj.** $\text{R}^{\text{2}}$ | **Overall model**  **p value** | **FDR-corrected p value** |
| --- | --- | --- | --- | --- | --- | --- | --- |
| **Global** | | | | | | | |
| Model 1 | 0.063 [-0.04 0.05] |  |  | 0.09 (1,23) | -0.039 | 0.766 | --- |
| Model 2 | 0.050 [-0.05 0.06] | -0.036 [-0.07 0.06] |  | 0.06 (2,22) | -0.085 | 0.946 | --- |
| Model 3 | 0.019 [-0.04 0.05] |  | 0.330 [-0.02 0.20] | 1.37 (2,22) | 0.030 | 0.274 | --- |
| Model 4 | -0.007 [-0.05 0.05] | -0.073 [-0.07 0.05] | 0.337 [-0.03 0.21] | 0.92 (3,21) | -0.011 | 0.450 | 0.450 |
| **Anterior region** | | | | | | | |
| Model 1 | 0.097 [-0.03 0.05] |  |  | 0.22 (1,23) | -0.034 | 0.645 | --- |
| Model 2 | 0.089 [-0.04 0.06] | -0.027 [-0.06 0.06] |  | 0.11 (2,22) | -0.080 | 0.895 | --- |
| Model 3 | 0.060 [-0.04 0.05] |  | 0.326 [-0.02 0.20] | 1.42 (2,22) | 0.034 | 0.263 | --- |
| Model 4 | 0.043 [-0.04 0.05] | -0.058 [-0.07 0.05] | 0.331 [-0.03 0.20] | 0.93 (3,21) | -0.009 | 0.444 | 0.450 |
| **Central region** | | | | | | | |
| Model 1 | 0.010 [-0.06 0.06] |  |  | <0.01 (1,23) | -0.043 | 0.964 | --- |
| Model 2 | -0.011 [-0.07 0.07] | -0.057 [-0.07 0.05] |  | 0.03 (2,22) | -0.088 | 0.968 | --- |
| Model 3 | -0.026 [-0.07 0.06] |  | 0.335 [-0.02 0.20] | 1.38 (2,22) | 0.031 | 0.273 | --- |
| Model 4 | -0.060 [-0.07 0.06] | -0.093 [-0.07 0.05] | 0.344 [-0.02 0.21] | 0.94 (3,21) | -0.007 | 0.437 | 0.450 |
| **Posterior region** | | | | | | | |
| Model 1 | -0.016 [-0.02 0.02] |  |  | 0.01 (1,23) | -0.043 | 0.938 | --- |
| Model 2 | -0.034 [-0.02 0.02] | -0.063 [-0.07 0.05] |  | 0.04 (2,22) | -0.087 | 0.958 | --- |
| Model 3 | -0.040 [-0.02 0.02] |  | 0.335 [-0.02 0.20] | 1.39 (2,22) | 0.031 | 0.270 | --- |
| Model 4 | -0.065 [-0.02 0.02] | -0.089 [-0.07 0.05] | 0.342 [-0.02 0.21] | 0.95 (3,21) | -0.006 | 0.434 | 0.450 |

**Table S6. Summary of omnibus test on multiple linear regression models testing group differences in peak alpha frequency (PAF), adjusted for the effects of age.** Two-group comparison (LC patient group and healthy pain-free control group) and 3-group comparison (moderate pain subgroup, severe pain subgroup and healthy pain-free control group) were tested on separate models. F statistic (F), degrees of freedom (df) and p values are reported for each independent variable in the model. Results relevant to each PAF extraction method (max-PAF and aperiodic-adjusted PAF) and each scalp region (global, anterior, central and posterior region) are presented in the table. * p < 0.05; ** p < 0.01.

| **Model** | | **Group** | | | **Age** | | |
| --- | --- | --- | --- | --- | --- | --- | --- |
|  |  | **F** | **df** | **p value** | **F** | **df** | **p value** |
| **Global** | | | | | | | |
| **Max-PAF models** | | | | | | | |
| 2-group model | | 1.56 | 1,59 | 0.217 | 0.24 | 1,59 | 0.626 |
| 3-group model |  | 3.43 | 2,54 | **0.040*** | 0.89 | 1,54 | 0.350 |
| **Aperiodic adjusted PAF-models** | | | | | | | |
| 2-group model | | 2.60 | 1,59 | 0.112 | 0.20 | 1,59 | 0.654 |
| 3-group model | | 3.39 | 2,54 | **0.041*** | 0.72 | 1,54 | 0.401 |
| **Anterior region** | | | | | | | |
| **Max-PAF models** | | | | | | | |
| 2-group model | | 3.62 | 1,59 | 0.062 | 0.01 | 1,59 | 0.925 |
| 3-group model | | 4.35 | 2,54 | **0.018*** | 0.20 | 1,54 | 0.658 |
| **Aperiodic adjusted PAF-models** | | | | | | | |
| 2-group model | | 3.88 | 1,59 | 0.054 | 0.25 | 1,59 | 0.619 |
| 3-group model | | 3.05 | 2,54 | 0.056 | 0.53 | 1,54 | 0.470 |
| **Central region** | | | | | | | |
| **Max-PAF models** | | | | | | | |
| 2-group model | | 0.90 | 1,59 | 0.345 | 0.18 | 1,59 | 0.674 |
| 3-group model | | 2.98 | 2,54 | 0.059 | 0.74 | 1,54 | 0.393 |
| **Aperiodic adjusted PAF-models** | | | | | | | |
| 2-group model | | 1.09 | 1,59 | 0.300 | 0.26 | 1,59 | 0.612 |
| 3-group model | | 2.56 | 2,54 | 0.087 | 0.89 | 1,54 | 0.349 |
| **Posterior region** | | | | | | | |
| **Max-PAF models** | | | | | | | |
| 2-group model | | 3.05 | 1,59 | 0.086 | 0.05 | 1,59 | 0.823 |
| 3-group model | | 5.07 | 2,54 | **0.010**** | 0.42 | 1,54 | 0.519 |
| **Aperiodic adjusted PAF-models** | | | | | | | |
| 2-group model | | 3.65 | 1,59 | 0.061 | 0.07 | 1,59 | 0.787 |
| 3-group model | | 5.07 | 2,54 | **0.010**** | 0.01 | 1,54 | 0.975 |

**Table S7. Post-hoc pairwise comparisons of peak alpha frequency (PAF) between groups, adjusted for the effects of age.** Only post-hoc pairwise comparisons from regression models showing a significant effect of group on omnibus test are presented in the table. Mean and standard deviation (SD) for each group, adjusted mean difference and 95% confidence intervals (CI), standard error (SE) and p values are reported for each pairwise comparison. Note that p values for the 3-group model (marked with †) are adjusted with Tukey method for multiple testing. * p < 0.05; ** p < 0.01.

| **Model** | **Comparison**  **(group 1 vs group 2)** | **Group 1 mean ± SD** | **Group 2 mean ± SD** | **Adjusted mean difference [95% CI]** | **SE** | **p value** |
| --- | --- | --- | --- | --- | --- | --- |
| **Global** | | | | | | |
| **Max-PAF models** | | | | | | |
| 3-group model | Moderate pain subgroup – healthy pain-free controls | 10.61 (±1.23) | 9.76 (±0.99) | 0.891 [0.01 1.78] | 0.37 | **0.049*** † |
|  | Severe pain subgroup – healthy pain-free controls | 9.71 (±0.99) | 9.76 (±0.99) | -0.095 [-0.88 0.69] | 0.32 | 0.954 † |
|  | Severe pain subgroup – moderate pain subgroup | 9.71 (±0.99) | 10.61 (±1.23) | -0.986 [-1.99 0.02] | 0.42 | 0.057 † |
| **Aperiodic-adjusted PAF models** | | | | | | |
| 3-group model | Moderate pain subgroup – healthy pain-free controls | 10.59 (±0.96) | 9.84 (±0.92) | 0.783 [0.02 1.55] | 0.32 | **0.044*** † |
|  | Severe pain subgroup – healthy pain-free controls | 9.83 (±0.79) | 9.84 (±0.92) | -0.037 [-0.71 0.64] | 0.28 | 0.990 † |
|  | Severe pain subgroup – moderate pain subgroup | 9.83 (±0.79) | 10.59 (±0.96) | -0.820 [-1.69 0.05] | 0.36 | 0.069 † |
| **Anterior region** | | | | | | |
| **max-PAF models** | | | | | | |
| 3-group model | Moderate pain subgroup – healthy pain-free controls | 10.55 (±1.25) | 9.53 (±0.95) | 1.039 [0.17 1.91] | 0.32 | **0.016*** † |
|  | Severe pain subgroup – healthy pain-free controls | 9.59 (±0.96) | 9.53 (±0.95) | 0.044 [-0.72 0.81] | 0.32 | 0.990 † |
|  | Severe pain subgroup – moderate pain subgroup | 9.59 (±0.96) | 10.55 (±1.25) | -0.996 [-1.99 -0.01] | 0.41 | **0.049*** † |
| **Posterior region** | | | | | | |
| **Max-PAF models** | | | | | | |
| 3-group model | Moderate pain subgroup – healthy pain-free controls | 10.98 (±1.16) | 9.91 (±1.10) | 1.118 [0.23 2.01] | 0.37 | **0.011*** † |
|  | Severe pain subgroup – healthy pain-free controls | 9.87 (±0.84) | 9.91 (±1.10) | -0.046 [-0.83 0.74] | 0.33 | 0.989 † |
|  | Severe pain subgroup – moderate pain subgroup | 9.87 (± 0.84) | 10.98 (±1.16) | -1.164 [-2.18 -0.15] | 0.42 | **0.021*** † |
| **Aperiodic-adjusted PAF models** | | | | | | |
| 3-group model | Moderate pain subgroup – healthy pain-free controls | 10.90 (±0.97) | 9.99 (±0.83) | 0.913 [0.19 1.63] | 0.30 | **0.009**** † |
|  | Severe pain subgroup – healthy pain-free controls | 9.99 (±0.74) | 9.99 (±0.83) | 0.001 [-0.63 0.63] | 0.26 | 1.000 † |
|  | Severe pain subgroup – moderate pain subgroup | 9.99 (±0.74) | 10.90 (±0.97) | -0.912 [-1.73 -0.10] | 0.34 | **0.025*** † |

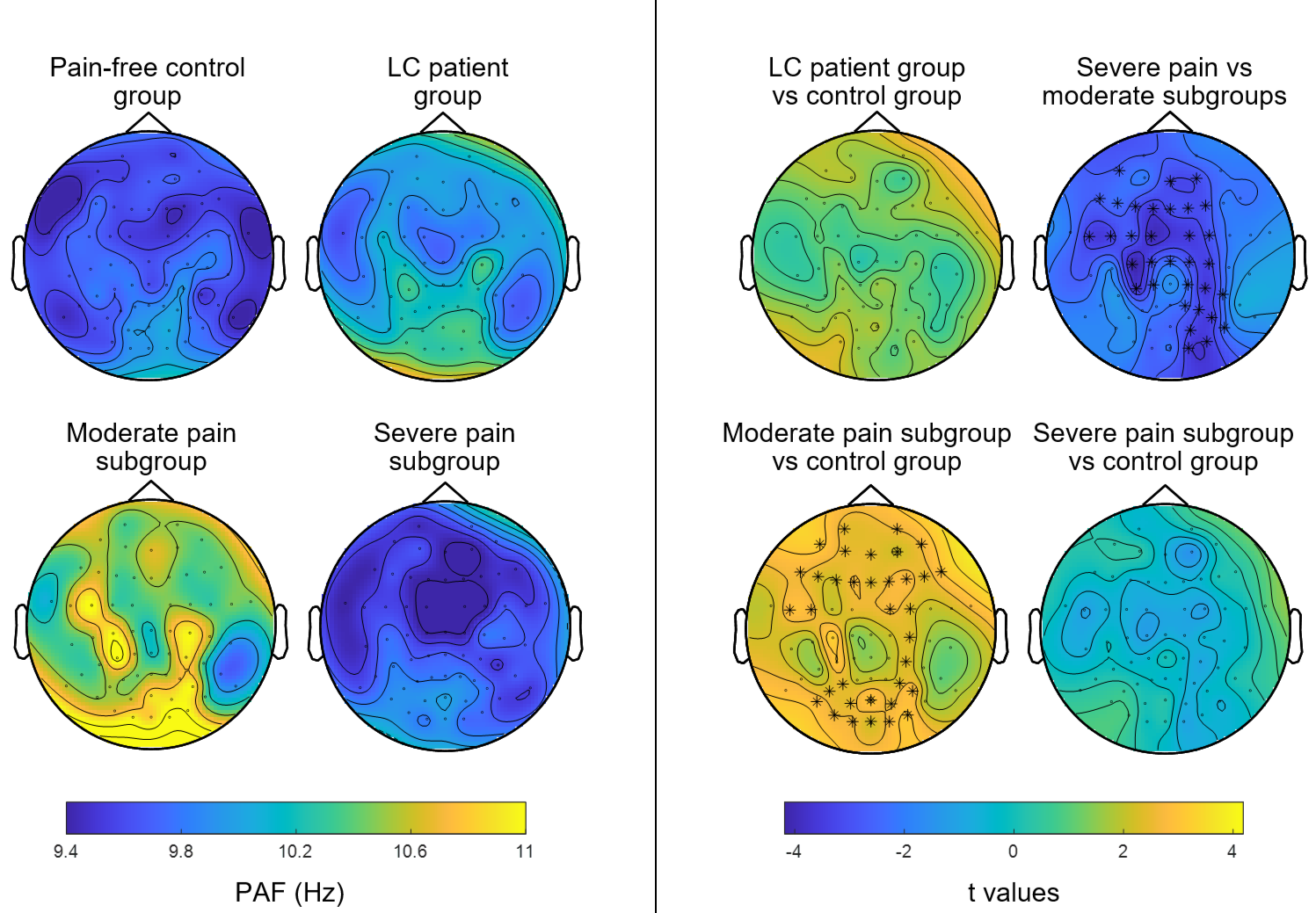

A

B

**Figure S1. Cluster-based permutation comparison of max-peak alpha frequency (max-PAF) between groups at channel level.** A) Topographical plots of PAF extracted for each electrode and averaged across participants within the healthy pain-free control group (n = 31), Long COVID (LC) patient group (n = 31), LC patient moderate pain subgroup (n = 11), and LC patient severe pain subgroup (n = 16). B) Channel-level comparison of PAF estimates between groups. Scalp maps represent topographical distribution of t-values for the comparison between groups, with warmer and colder colours representing increased and decreased PAF, respectively. Asterisks (*) denote electrode sites identified as part of significant clusters at significance level p < 0.05. Top left plot shows comparison between LC patient group and control group, with no significant clusters found. Top right plot shows comparison between severe pain and moderate pain LC patient subgroups, highlighting one significant negative cluster concentrated over frontal and central areas and extending into right-lateralised occipital-parietal electrode sites (p = 0.005, cluster statistic = -92.532). Bottom left panel shows comparison between patients reporting moderate pain and controls, highlighting one significant positive cluster covering bilateral frontal, right central and bilateral occipital-parietal regions (p = 0.005, cluster statistic = 87.134). Bottom right panel shows comparison between patients reporting severe pain and controls, with no significant clusters found. Data represented in the figure is derived from max-PAF estimates.

**Table S8. Summary of omnibus test on multiple linear regression models testing group differences in alpha band power, adjusted for the effects of age.** Two-group comparison (LC patient group and healthy pain-free control group) and 3-group comparison (moderate pain subgroup, severe pain subgroup and healthy pain-free control group) were tested on separate models. F statistic (F), degrees of freedom (df) and p values are reported for each independent variable in the model. Results relevant to global estimates and each tested scalp region (anterior, central and posterior region) are presented in the table.

| **Model** | | **Group** | | | **Age** | | |
| --- | --- | --- | --- | --- | --- | --- | --- |
|  |  | **F** | **df** | **p value** | **F** | **df** | **p value** |
| **Global** | | | | | | | |
| 2-group model | | 0.05 | 1,59 | 0.821 | 0.70 | 1,59 | 0.361 |
| 3-group model |  | 0.02 | 2,54 | 0.882 | 1.12 | 1,54 | 0.295 |
| **Anterior region** | | | | | | | |
| 2-group model | | 0.01 | 1,59 | 0.932 | 1.47 | 1,59 | 0.231 |
| 3-group model | | 0.15 | 2,54 | 0.863 | 1.83 | 1,54 | 0.182 |
| **Central region** | | | | | | | |
| 2-group model | | 0.09 | 1,59 | 0.760 | 0.21 | 1,59 | 0.645 |
| 3-group model | | 0.05 | 2,54 | 0.947 | 0.52 | 1,54 | 0.475 |
| **Posterior region** | | | | | | | |
| 2-group model | | 0.04 | 1,59 | 0.832 | 0.98 | 1,59 | 0.327 |
| 3-group model | | 0.11 | 2,54 | 0.899 | 1.02 | 1,54 | 0.316 |

**Exploratory analysis of association between posterior peak alpha frequency and pain severity accounting for use of centrally acting medication**

In order to determine whether use of centrally acting medication confounded the association between posterior PAF and pain severity, an additional exploratory multiple linear regression analysis was performed adjusting, in addition to previous confounders, for the potential confounding effects of medication status. Medication status was added as binary factor (taking centrally acting medication = 1, not taking centrally acting medication = 0) to the models. The models were specified as: *Pain severity* sc*ore ~ PAF + age + depression score + medication status.* The results show that medication status did not account for the association observed between posterior PAF and pain severity in LC patients with new-onset chronic pain.

**Table S9. Summary of multiple linear regression models on the association between posterior PAF and pain severity, controlling for the effects of age, depression and medication status.** Medication status was added as binary factor (taking centrally acting medication = 1, not taking centrally acting medication = 0) to the models. Models derived from max-PAF and aperiodic-adjusted PAF estimates are shown. Standardised β coefficients and standard errors for the independent variable and confounders, $\text{R}^{\text{2}}$, adjusted $\text{R}^{\text{2}}$, residual standard error, F statistic and degrees of freedom, and model p values are reported for each model. * p < 0.05; ** p < 0.01; *** p < 0.001.

|  | **Max-PAF** | **Aperiodic-adjusted PAF** |
| --- | --- | --- |
| **PAF** | **-0.618^**^** | **-0.593^**^** |
|  | (0.228) | (0.276) |
| **Age** | -0.032 | 0.063 |
|  | (0.022) | (0.023) |
| **Depression** | 0.209 | 0.277 |
|  | (0.046) | (0.047) |
| **Centrally acting drugs** | -0.060 | -0.046 |
|  | (0.511) | (0.522) |
| **Constant** | 14.764^***^ | 15.088^***^ |
|  | (2.672) | (2.924) |
| **R²** | 0.462 | 0.435 |
| **Adj. R²** | 0.355 | 0.322 |
| **Residual SE** | 1.158 | 1.186 |
| **F Statistic** | 4.30 (df=4;20) | 3.85 (df=4;20) |
| **Model p value** | **0.011*** | **0.018*** |
| *Note:* | ^*^p^**^p^***^p<0.001 | |

**Sensitivity analysis of group differences in peak alpha frequency including adjustment for the effects of depression**

Depression-adjusted between-group differences in PAF were tested with multiple linear regression models including a group factor and covariates age and depression. The group factor for the 2-group comparison consisted of 2-levels: LC patients vs controls; while the subgroup analysis models consisted of 3-levels: moderate pain group, severe pain group and control group. The models were specified as: *PAF ~ group + age + depression score.* Peak alpha frequency derived from the two estimation methods (max-PAF and aperiodic-adjusted PAF) for global and each tested scalp region (anterior, central, posterior) was entered as dependent variable in separate models. Each model was followed by an omnibus F-test of the coefficients which allowed to determine the overall effect of group, and subsequent Tukey’s adjusted post-hoc pairwise comparisons where group effects were significant (for details of omnibus test outputs and pairwise comparisons see Table S10 and Table S11, respectively).

Results of regression analysis testing depression-adjusted differences between groups were largely inconsistent across PAF estimation methods. When comparing overall LC patient group and controls, a significant main effect of group and depression was found for max-PAF estimates only, and not for aperiodic-adjusted PAF, at global and anterior region level; with post-hoc comparisons suggesting increased global and anterior max-PAF values in patients compared to controls. At subgroup level, a significant effect of group on PAF was found globally and in posterior scalp region, consistently for both max-PAF and aperiodic-adjusted PAF. Subsequent post-hoc pairwise comparisons showed increased global PAF in moderate pain subgroup compared to controls. In regard to posterior region, only the max-PAF model survived Tukey-adjusted pairwise comparison testing, highlighting significantly increased PAF in moderate pain subgroup compared to both severe pain subgroup and controls. A significant effect of group was also found in association with anterior and central max-PAF estimates, with moderate pain subgroup showing increased PAF compared to controls but no differences between pain severity subgroups.

**Table S10. Summary of omnibus test on multiple linear regression models testing group differences in peak alpha frequency (PAF), adjusted for the effects of age and depression.** Two-group comparison (LC patient group and healthy pain-free control group) and 3-group comparison (moderate pain subgroup, severe pain subgroup and healthy pain-free control group) were tested on separate models. F statistic (F), degrees of freedom (df) and p values are reported for each independent variable in the model. Results relevant to each PAF extraction method (max-PAF and aperiodic-adjusted PAF) and each scalp region (global, anterior, central and posterior region) are presented in the table. * p < 0.05.

| **Model** | | **Group** | | | **Age** | | | **Depression** | | |
| --- | --- | --- | --- | --- | --- | --- | --- | --- | --- | --- |
|  |  | **F** | **df** | **p value** | **F** | **df** | **p value** | **F** | **df** | **p value** |
| **Global** | | | | | | | | | | |
| **Max-PAF models** | | | | | | | | | | |
| 2-group model | | 4.49 | 1,52 | **0.039*** | 1.05 | 1,52 | 0.310 | 4.11 | 1,52 | **0.048*** |
| 3-group model |  | 4.42 | 2,51 | **0.017*** | 1.69 | 1,51 | 0.199 | 3.69 | 1,51 | 0.060 |
| **Aperiodic adjusted PAF-models** | | | | | | | | | | |
| 2-group model | | 2.80 | 1,52 | 0.1001 | 0.73 | 1,52 | 0.396 | 1.88 | 1,52 | 0.176 |
| 3-group model | | 3.32 | 2,51 | **0.044*** | 1.24 | 1,51 | 0.271 | 1.56 | 1,51 | 0.217 |
| **Anterior region** | | | | | | | | | | |
| **Max-PAF models** | | | | | | | | | | |
| 2-group model | | 7.11 | 1,52 | **0.010*** | 0.40 | 1,52 | 0.530 | 5.38 | 1,52 | **0.024*** |
| 3-group model | | 5.85 | 2,51 | **0.005*** | 0.82 | 1,51 | 0.370 | 4.93 | 1,51 | **0.031*** |
| **Aperiodic adjusted PAF-models** | | | | | | | | | | |
| 2-group model | | 2.95 | 1,52 | 0.092 | 0.66 | 1,52 | 0.418 | 1.36 | 1,52 | 0.249 |
| 3-group model | | 2.64 | 2,51 | 0.081 | 1.02 | 1,51 | 0.318 | 1.12 | 1,51 | 0.295 |
| **Central region** | | | | | | | | | | |
| **Max-PAF models** | | | | | | | | | | |
| 2-group model | | 3.78 | 1,52 | 0.057 | 0.81 | 1,52 | 0.372 | 3.86 | 1,52 | 0.055 |
| 3-group model | | 3.96 | 2,51 | **0.025*** | 1.37 | 1,51 | 0.248 | 3.45 | 1,51 | 0.069 |
| **Aperiodic adjusted PAF-models** | | | | | | | | | | |
| 2-group model | | 2.11 | 1,52 | 0.152 | 0.80 | 1,52 | 0.376 | 1.95 | 1,52 | 0.168 |
| 3-group model | | 2.81 | 2,51 | 0.069 | 1.30 | 1,51 | 0.260 | 1.63 | 1,51 | 0.207 |
| **Posterior region** | | | | | | | | | | |
| **Max-PAF models** | | | | | | | | | | |
| 2-group model | | 2.01 | 1,52 | 0.162 | 0.14 | 1,52 | 0.711 | 0.61 | 1,52 | 0.436 |
| 3-group model | | 4.43 | 2,51 | **0.017*** | 0.51 | 1,51 | 0.479 | 0.38 | 1,51 | 0.538 |
| **Aperiodic adjusted PAF-models** | | | | | | | | | | |
| 2-group model | | 0.96 | 1,52 | 0.3328 | 0.11 | 1,52 | 0.746 | 0.01 | 1,52 | 0.920 |
| 3-group model | | 3.90 | 2,51 | **0.026*** | 0.01 | 1,51 | 0.990 | 0.01 | 1,51 | 0.922 |

**Table S11. Post-hoc pairwise comparisons of peak alpha frequency (PAF) between groups, adjusted for the effects of age and depression.** Only post-hoc pairwise comparisons from regression models showing a significant effect of group on omnibus test are presented in the table. Mean and standard deviation (SD) for each group, adjusted mean difference and 95% confidence intervals (CI), standard error (SE) and p values are reported for each pairwise comparison. Note that p values for the 3-group model (marked with †) are adjusted with Tukey method for multiple testing. * p < 0.05; ** p < 0.01.

| **Model** | **Comparison**  **(group 1 vs group 2)** | **Group 1 mean ± SD** | **Group 2 mean ± SD** | **Adjusted mean difference [95% CI]** | **SE** | **p value** |
| --- | --- | --- | --- | --- | --- | --- |
| **Global** | | | | | | |
| **Max-PAF models** | | | | | | |
| 2-group model | All patients –  healthy pain-free controls | 10.11 (±1.15) | 9.76 (±0.99) | 0.950 [0.05 1.85] | 0.45 | **0.039*** |
| 3-group model | Moderate pain subgroup – healthy pain-free controls | 10.61 (±1.23) | 9.76 (±0.99) | 1.445 [0.24 2.65] | 0.50 | **0.015*** † |
|  | Severe pain subgroup – healthy pain-free controls | 9.71 (±0.99) | 9.76 (±0.99) | 0.576 [-0.57 1.72] | 0.47 | 0.449 † |
|  | Severe pain subgroup – moderate pain subgroup | 9.71 (±0.99) | 10.61 (±1.23) | -0.869 [-1.91 0.17] | 0.43 | 0.118 † |
| **Aperiodic-adjusted PAF models** | | | | | | |
| 3-group model | Moderate pain subgroup – healthy pain-free controls | 10.59 (±0.96) | 9.84 (±0.92) | 1.070 [0.01 2.13] | 0.44 | **0.047*** † |
|  | Severe pain subgroup – healthy pain-free controls | 9.83 (±0.79) | 9.84 (±0.92) | 0.344 [-0.66 1.35] | 0.41 | 0.687 † |
|  | Severe pain subgroup – moderate pain subgroup | 9.83 (±0.79) | 10.59 (±0.96) | -0.726 [-1.64 0.19] | 0.38 | 0.143 † |
| **Anterior region** | | | | | | |
| **max-PAF models** | | | | | | |
| 2-group model | All patients –  healthy pain-free controls | 10.06 (±1.22) | 9.53 (±0.95) | 1.160 [0.29 2.03] | 0.43 | **0.010*** |
| 3-group model | Moderate pain subgroup – healthy pain-free controls | 10.55 (±1.25) | 9.53 (±0.95) | 1.640 [0.47 2.81] | 0.48 | **0.004**** † |
|  | Severe pain subgroup – healthy pain-free controls | 9.59 (±0.96) | 9.53 (±0.95) | 0.791 [-0.31 1.90] | 0.46 | 0.204 † |
|  | Severe pain subgroup – moderate pain subgroup | 9.59 (±0.96) | 10.55 (±1.25) | -0.849 [-1.85 0.16] | 0.42 | 0.113 † |
| **Central region** | | | | | | |
| **Max-PAF models** | | | | | | |
| 3-group model | Moderate pain subgroup – healthy pain-free controls | 10.59 (±1.20) | 9.81 (±1.04) | 1.394 [0.15 2.63] | 0.51 | **0.024*** † |
|  | Severe pain subgroup – healthy pain-free controls | 9.70 (±1.00) | 9.81 (±1.04) | 0.517 [-0.66 1.69] | 0.49 | 0.541 † |
|  | Severe pain subgroup – moderate pain subgroup | 9.70 (±1.00) | 10.59 (±1.20) | -0.876 [-1.94 0.19] | 0.44 | 0.127 † |
| **Posterior region** | | | | | | |
| **Max-PAF models** | | | | | | |
| 3-group model | Moderate pain subgroup – healthy pain-free controls | 10.98 (±1.16) | 9.91 (±1.10) | 0.747 [0.01 1.48] | 0.30 | **0.045*** † |
|  | Severe pain subgroup – healthy pain-free controls | 9.87 (±0.84) | 9.91 (±1.10) | -0.449 [-1.52 0.62] | 0.44 | 0.571 † |
|  | Severe pain subgroup – moderate pain subgroup | 9.87 (± 0.84) | 10.98 (±1.16) | -1.196 [-2.32 -0.07] | 0.47 | **0.035*** † |
| **Aperiodic-adjusted PAF models** | | | | | | |
| 3-group model | Moderate pain subgroup – healthy pain-free controls | 10.90 (±0.97) | 9.99 (±0.83) | 0.584 [-0.01 1.18] | 0.25 | 0.057 † |
|  | Severe pain subgroup – healthy pain-free controls | 9.99 (±0.74) | 9.99 (±0.83) | -0.207 [-1.08 0.67] | 0.36 | 0.836 † |
|  | Severe pain subgroup – moderate pain subgroup | 9.99 (±0.74) | 10.90 (±0.97) | -0.791 [-1.71 0.13] | 0.38 | 0.106 † |

**Exploratory analysis of differences in delta, theta and beta spectral power between patients and controls**

**Table S12. Summary of multiple linear regression models on the association between absolute spectral power of canonical EEG frequency bands and pain severity.** Model 1 represents results of simple linear regression between dependent variable (pain severity) and independent variable (absolute spectral power of each canonical EEG frequency band: delta, theta and beta). Models 2 and 3 outline the effects of introducing each confounding variable (age and depression, respectively); and Model 4 includes confounding effects of both age and depression. Standardised β coefficients and 95% confidence intervals (CI) for each independent variable, F statistic and degrees of freedom, adjusted $\text{R}^{\text{2}}$ and raw overall model p values are reported for each combination of EEG frequency band (delta, theta and beta), scalp region (global, anterior, central and posterior region), and model (model 1 to model 4).

|  | **Standardised β coefficient for spectral power [95% CI]** | **Standardised β coefficient for age [95% CI]** | **Standardised β coefficient for depression**  **[95% CI]** | **F statistic (degrees of freedom)** | **Adj.** $\text{R}^{\text{2}}$ | **Overall model p value** |
| --- | --- | --- | --- | --- | --- | --- |
| **Global** | | | | | | |
| **Delta band power** | | | | | | |
| Model 1 | <0.001 [-0.26 0.26] |  |  | <0.01 (1,23) | -0.043 | 0.999 |
| Model 2 | -0.045 [-0.35 0.30] | -0.079 [-0.08 0.06] |  | 0.05 (2,22) | -0.086 | 0.955 |
| Model 3 | -0.049 [-0.28 0.23] |  | 0.340 [-0.02 0.20] | 1.40 (2,22) | 0.032 | 0.267 |
| Model 4 | -0.139 [-0.40 0.24] | -0.151 [-0.09 0.05] | 0.361 [-0.02 0.21] | 1.03 (3,21) | 0.003 | 0.401 |
| **Theta band power** | | | | | | |
| Model 1 | 0.342 [-0.01 0.13] |  |  | 3.04 (1,23) | 0.078 | 0.094 |
| Model 2 | 0.340 [-0.01 0.13] | -0.011 [-0.06 0.05] |  | 1.46 (2,22) | 0.037 | 0.255 |
| Model 3 | 0.257 [-0.03 0.12] |  | 0.243 [-0.05 0.18] | 2.23 (2,22) | 0.093 | 0.131 |
| Model 4 | 0.251 [-0.03 0.12] | -0.035 [-0.06 0.05] | 0.247 [-0.05 0.19] | 1.43 (3,21) | 0.051 | 0.262 |
| **Beta band power** | | | | | | |
| Model 1 | 0.071 [-0.10 0.14] |  |  | 0.12 (1,23) | -0.038 | 0.735 |
| Model 2 | 0.065 [-0.11 0.14] | -0.044 [-0.06 0.05] |  | 0.08 (2,22) | -0.083 | 0.926 |
| Model 3 | 0.159 [-0.07 0.16] |  | 0.371 [-0.01 0.21] | 1.71 (2,22) | 0.056 | 0.204 |
| Model 4 | 0.152 [-0.08 0.17] | -0.050 [-0.06 0.05] | 0.372 [-0.02 0.22] | 1.11 (3,21) | 0.014 | 0.366 |
| **Anterior region** | | | | | | |
| **Delta band power** | | | | | | |
| Model 1 | 0.050 [-0.31 0.40] |  |  | 0.06 (1,23) | -0.041 | 0.810 |
| Model 2 | 0.028 [-0.44 0.49] | -0.035 [-0.08 0.07] |  | 0.04 (2,22) | -0.087 | 0.964 |
| Model 3 | -0.039 [-0.39 0.32] |  | 0.343 [-0.02 0.21] | 1.39 (2,22) | 0.031 | 0.270 |
| Model 4 | -0.155 [-0.61 0.36] | -0.169 [-0.10 0.05] | 0.382 [-0.02 0.23] | 1.03 (3,21) | 0.004 | 0.400 |
| **Theta band power** | | | | | | |
| Model 1 | 0.347 [-0.01 0.11] |  |  | 3.15 (1,23) | 0.082 | 0.089 |
| Model 2 | 0.345 [-0.01 0.11] | -0.027 [-0.06 0.05] |  | 1.52 (2,22) | 0.041 | 0.241 |
| Model 3 | 0.263 [-0.02 0.10] |  | 0.240 [-0.05 0.18] | 2.28 (2,22) | 0.096 | 0.126 |
| Model 4 | 0.258 [-0.03 0.10] | -0.046 [-0.06 0.05] | 0.245 [-0.05 0.18] | 1.47 (3,21) | 0.055 | 0.251 |
| **Beta band power** | | | | | | |
| Model 1 | 0.107 [-0.09 0.15] |  |  | 0.26 (1,23) | -0.032 | 0.612 |
| Model 2 | 0.100 [-0.10 0.16] | -0.035 [-0.06 0.05] |  | 0.14 (2,22) | -0.077 | 0.870 |
| Model 3 | 0.200 [-0.06 0.18] |  | 0.382 [-0.01 0.22] | 1.91 (2,22) | 0.071 | 0.171 |
| Model 4 | 0.193 [-0.07 0.18] | -0.039 [-0.06 0.05] | 0.382 [-0.01 0.22] | 1.23 (3,21) | 0.028 | 0.323 |
| **Central region** | | | | | | |
| **Delta band power** | | | | | | |
| Model 1 | 0.030 [-0.26 0.29] |  |  | 0.02 (1,23) | -0.043 | 0.888 |
| Model 2 | 0.004 [-0.32 0.33] | -0.051 [-0.07 0.06] |  | 0.03 (2,22) | -0.088 | 0.969 |
| Model 3 | -0.012 [-0.28 0.26] |  | 0.334 [-0.02 0.20] | 1.37 (2,22) | 0.030 | 0.275 |
| Model 4 | -0.066 [-0.36 0.28] | -0.104 [-0.08 0.05] | 0.346 [-0.02 0.21] | 0.94 (3,21) | -0.007 | 0.437 |
| **Theta band power** | | | | | | |
| Model 1 | 0.329 [-0.01 0.14] |  |  | 2.80 (1,23) | 0.070 | 0.108 |
| Model 2 | 0.328 [-0.02 0.14] | -0.015 [-0.06 0.05] |  | 1.34 (2,22) | 0.028 | 0.282 |
| Model 3 | 0.250 [-0.03 0.13] |  | 0.255 [-0.05 0.18] | 2.21 (2,22) | 0.091 | 0.134 |
| Model 4 | 0.244 [-0.04 0.13] | -0.038 [-0.06 0.05] | 0.258 [-0.05 0.19] | 1.42 (3,21) | 0.050 | 0.265 |
| **Beta band power** | | | | | | |
| Model 1 | 0.037 [-0.11 0.13] |  |  | 0.03 (1,23) | -0.042 | 0.861 |
| Model 2 | 0.035 [-0.11 0.13] | -0.052 [-0.07 0.05] |  | 0.04 (2,22) | -0.086 | 0.956 |
| Model 3 | 0.138 [-0.08 0.16] |  | 0.370 [-0.01 0.21] | 1.62 (2,22) | 0.049 | 0.221 |
| Model 4 | 0.136 [-0.08 0.16] | -0.069 [-0.06 0.05] | 0.373 [-0.02 0.22] | 1.07 (3,21) | 0.009 | 0.382 |
| **Posterior region** | | | | | | |
| **Delta band power** | | | | | | |
| Model 1 | -0.018 [-0.24 0.22] |  |  | 0.01 (1,23) | -0.043 | 0.932 |
| Model 2 | -0.080 [-0.34 0.26] | -0.102 [-0.09 0.06] |  | 0.08 (2,22) | -0.083 | 0.927 |
| Model 3 | -0.076 [-0.27 0.19] |  | 0.345 [-0.02 0.21] | 1.45 (2,22) | 0.036 | 0.257 |
| Model 4 | -0.200 [-0.40 0.19] | -0.195 [-0.10 0.05] | 0.377 [-0.02 0.22] | 1.13 (3,21) | 0.016 | 0.359 |
| **Theta band power** | | | | | | |
| Model 1 | 0.339 [-0.01 0.10] |  |  | 2.99 (1,23) | 0.077 | 0.097 |
| Model 2 | 0.338 [-0.01 0.10] | -0.011 [-0.06 0.05] |  | 1.43 (2,22) | 0.035 | 0.260 |
| Model 3 | 0.256 [-0.02 0.09] |  | 0.246 [-0.05 0.18] | 2.23 (2,22) | 0.093 | 0.131 |
| Model 4 | 0.250 [-0.03 0.10] | -0.035 [-0.06 0.05] | 0.250 [-0.05 0.19] | 1.43 (3,21) | 0.051 | 0.261 |
| **Beta band power** | | | | | | |
| Model 1 | 0.023 [-0.08 0.09] |  |  | 0.01 (1,23) | -0.043 | 0.911 |
| Model 2 | 0.001 [-0.09 0.09] | -0.050 [-0.07 0.05] |  | 0.03 (2,22) | -0.088 | 0.968 |
| Model 3 | 0.083 [-0.07 0.10] |  | 0.347 [-0.02 0.21] | 1.46 (2,22) | 0.037 | 0.253 |
| Model 4 | 0.069 [-0.08 0.10] | -0.053 [-0.07 0.05] | 0.347 [-0.02 0.21] | 0.95 (3,21) | -0.006 | 0.432 |

**Table S13. Summary of omnibus test on multiple linear regression models testing group differences in absolute band power of canonical EEG frequency bands, adjusted for the effects of age.** Two-group comparison (LC patient group and healthy pain-free control group) and 3-group comparison (moderate pain subgroup, severe pain subgroup and healthy pain-free control group) were tested on separate models. F statistic (F), degrees of freedom (df) and p values are reported for each independent variable in the model. Results relevant to each EEG frequency band (delta, theta and beta) and each tested scalp region (global, anterior, central and posterior region) are presented in the table.

| **Model** | | **Group** | | | **Age** | | |
| --- | --- | --- | --- | --- | --- | --- | --- |
|  |  | **F** | **df** | **p value** | **F** | **df** | **p value** |
| **Global** | | | | | | | |
| **Delta band power** | | | | | | | |
| 2-group model | | 0.03 | 1,59 | 0.870 | 1.71 | 1,59 | 0.197 |
| 3-group model |  | 0.01 | 2,54 | 0.988 | 1.73 | 1,54 | 0.194 |
| **Theta band power** | | | | | | | |
| 2-group model | | 0.08 | 1,59 | 0.782 | 1.10 | 1,59 | 0.298 |
| 3-group model | | 0.74 | 2,54 | 0.482 | 1.14 | 1,54 | 0.289 |
| **Beta band power** | |  |  |  |  |  |  |
| 2-group model | | 0.01 | 1,59 | 0.937 | 1.50 | 1,59 | 0.226 |
| 3-group model | | 0.16 | 2,54 | 0.851 | 1.32 | 1,54 | 0.256 |
| **Anterior region** | | | | | | | |
| **Delta band power** | | | | | | | |
| 2-group model | | 0.73 | 1,59 | 0.396 | 2.98 | 1,59 | 0.089 |
| 3-group model | | 0.31 | 2,54 | 0.734 | 2.97 | 1,54 | 0.090 |
| **Theta band power** | | | | | | | |
| 2-group model | | 0.02 | 1,59 | 0.893 | 1.60 | 1,59 | 0.211 |
| 3-group model | | 0.96 | 2,54 | 0.388 | 1.69 | 1,54 | 0.199 |
| **Beta band power** | |  |  |  |  |  |  |
| 2-group model | | <0.01 | 1,59 | 0.982 | 0.98 | 1,59 | 0.325 |
| 3-group model | | 0.34 | 2,54 | 0.710 | 0.86 | 1,54 | 0.359 |
| **Central region** | | | | | | | |
| **Delta band power** | | | | | | | |
| 2-group model | | 0.09 | 1,59 | 0.762 | 0.46 | 1,59 | 0.501 |
| 3-group model | | 0.16 | 2,54 | 0.853 | 0.44 | 1,54 | 0.511 |
| **Theta band power** | | | | | | | |
| 2-group model | | 0.08 | 1,59 | 0.773 | 0.74 | 1,59 | 0.393 |
| 3-group model | | 0.60 | 2,54 | 0.553 | 0.85 | 1,54 | 0.359 |
| **Beta band power** | |  |  |  |  |  |  |
| 2-group model | | 0.19 | 1,59 | 0.660 | 2.96 | 1,59 | 0.091 |
| 3-group model | | 0.26 | 2,54 | 0.772 | 2.60 | 1,54 | 0.113 |
| **Posterior region** | | | | | | | |
| **Delta band power** | | | | | | | |
| 2-group model | | 0.52 | 1,59 | 0.475 | 0.93 | 1,59 | 0.338 |
| 3-group model | | 0.10 | 2,54 | 0.903 | 0.84 | 1,54 | 0.362 |
| **Theta band power** | | | | | | | |
| 2-group model | | 0.03 | 1,59 | 0.853 | 0.43 | 1,59 | 0.514 |
| 3-group model | | 0.59 | 2,54 | 0.558 | 0.36 | 1,54 | 0.556 |
| **Beta band power** | |  |  |  |  |  |  |
| 2-group model | | <0.01 | 1,59 | 0.976 | 0.42 | 1,59 | 0.518 |
| 3-group model | | 0.05 | 2,54 | 0.952 | 0.40 | 1,54 | 0.527 |
